# High retention rates of custom 3D printed titanium implants in complex pelvic reconstruction, a report on 106 consecutive cases over 10 years

**DOI:** 10.1101/2024.10.02.24313775

**Authors:** Richard Boyle, Corey Scholes, Daniel Franks, Amish Lodhia, Meredith Harrison-Brown, Milad Ebrahimi, Maurice Guzman, Paul Stalley

## Abstract

**Background:** Access to custom 3D printed pelvic implants (3DPI) is improving for application in both arthroplasty revision and tumour reconstruction. There is limited evidence regarding the safety and outcomes of such implants for large bony defects of the pelvis.

**Purpose:** To report the incidence of complications, patient mortality and implant survival following pelvic reconstruction using custom 3Dprinted prostheses in the setting of extensive pelvic bone defects following pelvic tumour resection or failure of total hip arthroplasty (THA)

**Methods:** Patients who underwent reconstruction with a custom 3D printed pelvic prosthesis (3DPI) were identified from our clinical outcomes registry (Complex Reconstruction and Sarcoma Surgical Outcomes Registry; ANZCTRN 12621001421820). Indications for surgery, adverse events, reoperations and rates and modes of failure were recorded. Kaplan-Meier and multistate survival curves were generated for cumulative survival based on indication.

**Results:** One hundred and six procedures were completed(RevisionTHA = 33; TumourPelvis = 73) with a median follow up of 4.1 years, ranging from 0.6 to 10 years. Acetabular loosening was the most frequent indication for the RevisionTHA cohort, while indications for tumour varied across primary presentations, metastases and failures of previous resection/reconstruction. Intraoperative complications were observed in 4.1% (95%CI 1.1 - 12.3) of TumourPelvis cases. Overall implant retention was 96% (90 - 99). No mortality events were observed in the RevisionTHA cohort, with 5-year patient survival 79% (70-90) in the TumourPelvis cohort. Procedure-survival free from periprosthetic infection was 86% (74-100) in the RevisionTHA cohort and 85% (76 - 95) in the TumourPelvisCohort. Modelling adverse events using multistate survival models in both cohorts revealed complex time-varying presentation of adverse events, with a significant burden of reoperations and local tumour recurrence in the TumourPelvis cohort.

**Conclusion:** 3DPIs are a safe and viable option for complex reconstruction of the pelvis across a range of oncological and non-oncological indications. The initial results of the present study provide important information to aid in counselling patients about such procedures and allocating healthcare resources for ongoing care. Further work is required to document functional and biomechanical outcomes in these patient populations.

## Introduction

Extensive bone loss in the pelvis due to tumour resection or failed arthroplasty poses a significant challenge for surgical reconstruction. The goals of reconstruction in this situation are restoration of functional bony and soft tissue anatomy, obtaining primary and secondary fixation of the construct to remaining bone, as well as optimising adjacent joint kinematics and fatigue resistance of the construct. The challenge in large pelvic reconstruction is achieving these goals in the context of unique patient anatomy and highly variable defect size and shape. Many techniques have been described to restore function after pelvic bone loss (Pandey et al., 2024), which rely on standard “off the shelf” prostheses and often fail to achieve all the goals of reconstruction (Wu et al., 2023).

Custom made prostheses for pelvic reconstruction (3DPI), applying three-dimensional printing (additive manufacturing) techniques to titanium and other suitable materials, offer a bespoke solution to pelvic bone defects. The advantages of 3DPI go beyond the ability to customise the size and shape of the prosthesis, but also in the design process itself, which necessitates computer modelling of the individual’s relevant anatomy, pathology, defect and existing hardware. Computer aided design of the implant allows accurate planning of geometry of the bone implant interface, fixation elements, screw trajectory, implant strength, hip centre and cup orientation. The process of 3D printing is relatively fast and accurate, and enables utilisation of osseointegrative microstructure on bone interface surfaces to maximise osseointegration and secondary fixation. Finally, the process also allows for manufacture of customised resection jigs for both tumour resection and revision arthroplasty, which accurately prepares the bone surfaces to accept the implant, improving accuracy of implant position and decreasing operative time. Many of these benefits hinge on nuances of the design process.

Work of our group and others has shown favourable early results of 3DPI in both pelvic tumour resections(Broekhuis et al., 2022), and failed total hip replacement(Kieser et al., 2018) However, the evidence regarding their efficacy and safety remains limited and we have shown previously that major pelvic tumour resection with 3DPI reconstruction poses a considerable challenge for anaesthetic management and has a high risk of complications and other adverse events after implantation(Chua et al., 2019). More broadly, there is also concern about increased infection risk due to the large surface area of titanium-based 3DPI (Sui et al., 2022), and the presence of incompletely sintered residual titanium particles (Tang et al., 2023).

3DPI to treat pelvic bone defects in revision arthroplasty and ortho oncology is becoming more accessible, but small case numbers have limited the accumulation of experience or development of a robust design process for pelvic prostheses. To address the apprehension regarding the broader uptake of this technology without sufficient knowledge of clinical tolerance and implant efficacy, this study reports the complication rate, mortality and implant survival following 3DPI pelvic reconstruction with 3D the design process and surgical technique developed in our unit.

## Methods

The present analysis is a retrospective cohort study embedded within a prospective clinical quality registry (Complex Reconstruction and Sarcoma Surgical Outcomes Registry; ANZCTRN 12621001421820). The study is reported based on the STROBE guidelines (von Elm et al., 2007), with a checklist included as a supplementary file (Supplementary 1). Ethical approval for the COMPRESSOR Registry was obtained from the Local Health District (RPA Zone) Human Research Ethics Committee (Ref 2021/ETH00634), which included a waiver of consent approval for chart review. The dataset represents all records captured for a surgical group within Sydney, Australia using the implant of interest from its market introduction to the date of analysis. All reviewed charts from the operating surgeons practice records (electronic medical record, paper files where required) were entered manually into the database and the present analysis draws data from the live database tables directly. The review was completed 1-Aug-2023 and the dataset was last retrieved 28-Aug-2024.

### Patient and case selection

Patients were included for analysis if they met the following criteria: aged 16 or over on the date of surgery and received a custom 3DPI for pelvic reconstruction for any indication. A consecutive series was identified through a common label for procedures involving the implant type of interest and retrieved from the database. Chart review was performed on practice treatment records of the participating surgeons and the data aligned to the data schema of the clinical registry. The analysis sample included two cohorts within the registry: TumourPelvis and Revision Total Hip Arthroplasty (RevisionTHA). The TumourPelvis cohort included primary tumour of bone, metastatic disease and revision of previous tumour reconstruction. The RevisionTHA cohort included cases presenting with a failed hip prosthesis in-situ in which the size of the acetabular defect (Type 3) (Paprosky et al., 1994) was deemed unsuitable for standard revision components.

### Prosthesis Design

Implants were designed and manufactured by OSSIS Limited (Christchurch, New Zealand). Data from computed tomography (CT) and magnetic resonance imaging (MRI) scans of the pelvis were obtained and modelled (Mimics v. 23; Materialise, Belgium), to identify bone margins and defects. Key neural structures were segmented and modelled where required to ensure sufficient clearance during and after implantation. When hip replacement was necessary, the hip centre coordinates were established by matching to the contralateral hip. Acetabular orientation was set at 40 degrees abduction, 20 degrees anteversion. A hemispherical acetabular shell with no internal surface geometry but a roughened surface was modelled to accept a cemented cup, with the capability to change orientation intraoperatively.

Implant to bone interface geometry and cortical flanges were planned to provide intrinsic stability based on the modelled defect. Screw trajectory was planned to intersect with the highest quality bone on the basis of CT, avoid neural structures and to cross the sacroiliac joint where adequate iliac fixation was not achievable. Screw trajectory also considered surgical access to achieve the necessary drill and driver orientation based on the approach chosen. Inclusion of anterior fixation was dependent on the amount of pubic bone remaining for fixation, or left unfixed (leaving the pelvic ring “open”). The body of the prosthesis was designed to adequately transmit load between the fixation elements and the acetabular shell whilst minimising prosthesis volume and surface area. Ingrowth surfaces were designed at the bone interface surfaces and an open mesh included to fill non load-bearing voids between the bone and implant.

### Surgical technique

All operations were performed by one of the senior authors of this paper (RB/PS/DF/MG) as previously described(Broekhuis et al., 2022; Chua et al., 2019). Patients were positioned supine with use of an extended iliofemoral approach. Suitable exposure of the pelvic sidewall was achieved by reflecting the gluteal muscles laterally, with anterior superior iliac crest osteotomy used where possible. Where superior pubic ramus or pubic symphysis resection was necessary, a second Pfannenstiel type inline incision was utilised, with care to leave an adequate skin bridge between the two incisions. For posterior sacroiliac resections the patients were repositioned prone after closure of the anterior wound, with use of a curvilinear incision vertically along the spine and laterally into the buttock parallel to gluteal muscle fibres. Wounds were closed over suction drains. All patients were admitted to the intensive care unit (ICU) immediately following surgery, and all received chemical thromboprophylaxis for 6 weeks and a prophylactic antibiotic regimen (intravenous vancomycin and cefepime followed by oral cephalexin). Patients remained on bed rest until muscle control was regained, with rehabilitation followed under physiotherapist supervision and guidance pre and post-discharge.

For Enneking P2 resections (periacetabular) where femoral prosthetic reconstruction was required, either standard or revision arthroplasty was performed using a cemented femoral stem or megaprosthetic femoral replacement (Exeter or Global Modular Replacement System, Stryker, Mahwah, New Jersey, United States, or Taperfit, Corin). On the acetabular side, a semi- constrained polyethylene Snap-Fit cup (Bioimpianti, Milan, Italy) or dual-mobility component Trinity cup (Corin, Circester, UK), of fully constrained cemented component (Stryker Trident) was cemented into the acetabular shell of the pelvic prosthesis using PMMA cement depending on intraoperative assessment of stability.

### Outcomes

A full description of the variables collected for the analysis is included in Supplementary 2. The outcomes of interest for this study were complications, revisions, reoperations, implant survival and patient mortality. Complications were defined as any deviation from the expected course of treatment and recovery. Revision of the index procedure was defined as any subsequent operation involving the replacement or reseating of the 3DPI or the associated acetabular liner or femoral stem as per the AOANJRR (Australian Orthopaedic Association National Joint Replacement Registry). Reoperations were defined as any surgical intervention on the index area of treatment not meeting the criteria for a revision. Implant-related failure was defined as any failure due to aseptic loosening of the implant; implant or fixation breakage. Failure mechanisms considered not related to the implant were tumour recurrence; soft tissue failure; hip joint dislocation or failure of the femoral component. Periprosthetic infection was considered separately.

### Data and Statistical Analysis

Data pre-processing and analysis was conducted in the R platform (R Core Team, 2024) with specialised packages used where required, as described in Supplementary 2. Confidence intervals were included for descriptive statistics (patient characteristics and complications), while follow-up durations were described with median and interquartile range. Text processing tools were used to extract diagnoses, complications and reoperations from registry records. Regular expressions (regex) were constructed to extract complications as described by (Healy et al., 2016) and modes of failure for tumour endoprostheses according to the Henderson system(Henderson et al., 2014). The dataset was subset multiple times corresponding to each type of complication and the dataset modified to incorporate a status column (complication present or absent) and the duration between the event date (or follow up date if complication absent). Kaplan-Meier survival curves (single event per record) were generated for assessment of survival from complications, mortality and amputation|implant removal. The survival analysis was supplemented with a multi-state model, whereby each patient can present with multiple adverse events in sequence (sometimes of the same type) which can violate some assumptions of the Kaplan-Meier analysis (Thenmozhi et al., 2019) and can lead to bias where competing events are censored from the patient pool (Coemans et al., 2022).

## Results

A sample of 106 index cases meeting the inclusion criteria were included for analysis (Figure 1). Of note was a low abandonment incidence (N = 3, 2.8%, 95%CI 0.6 - 8.0%) Two cases were abandoned due to intraoperative blood loss and one case due to tumour progression.

**Figure 1:**
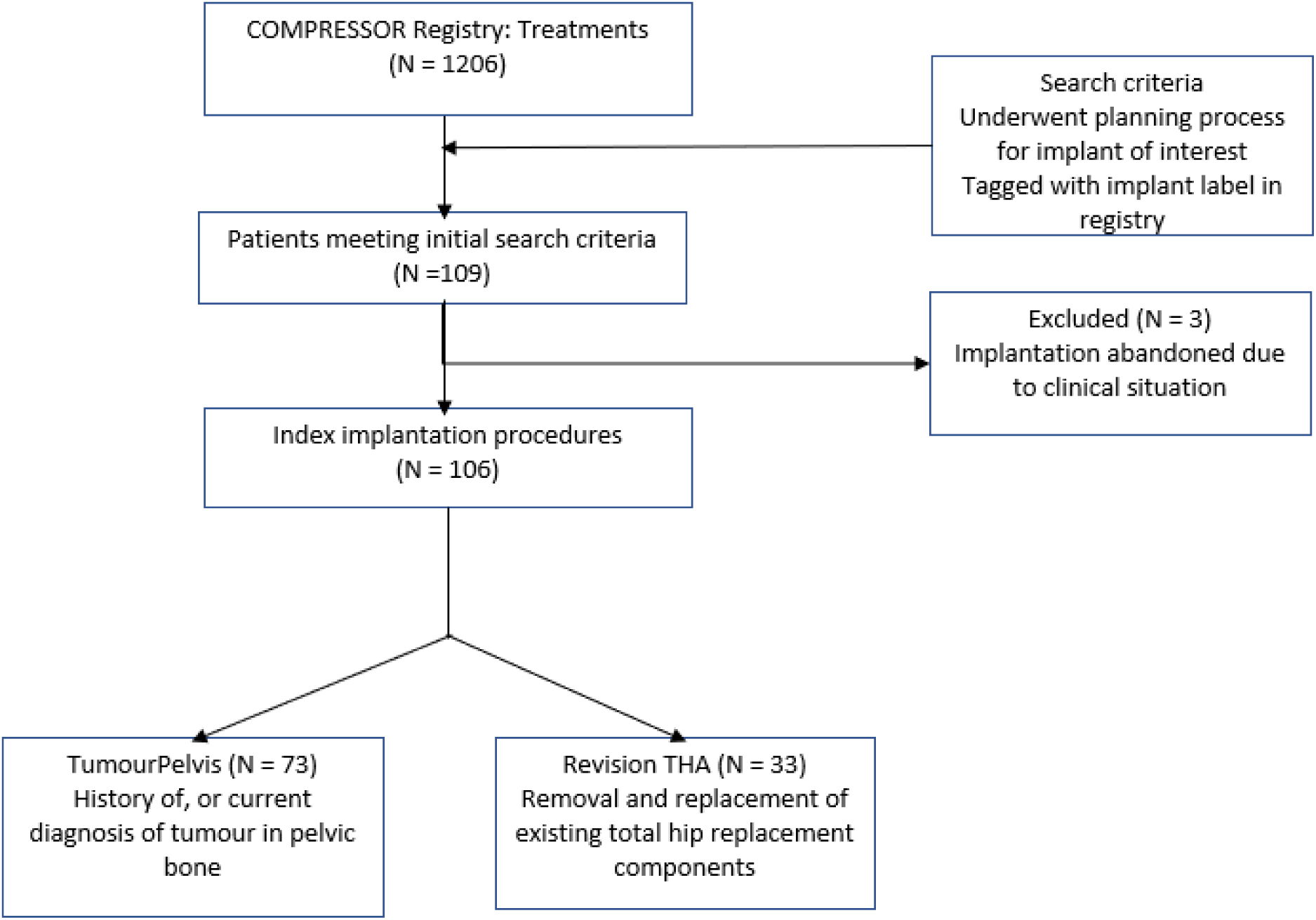
STROBE flow chart for pelvic reconstructions added to COMPRESSOR.

### Patient characteristics

Patients included in the series were aged 55 years on average and evenly distributed between males and females and sides of the pelvis, with two surgeons performing the majority of the cases (Table 1). The patient demographics did vary when considering the RevisionTHA and TumourPelvis cohorts (Table 1). The indications for the TumourPelvis cohort were sarcoma, metastasis to the pelvis and other tumours of the pelvis (Table 2a). The RevisionTHA cohort presented with several overlapping indications (Table 2b), with aseptic loosening of the acetabulum common to all cases.

**Table 1:** Summary of patient characteristics receiving implant of interest - separated by registry cohort.

| Characteristic | Overall<br>N = 106 | 95% CI<sup>1</sup> | RevisTHA<sup>1</sup> N<br>= 33 | 95% CI<sup>2</sup> | TumourPelvis*<br>N = 73 | 95% CI<sup>1</sup> |
| --- | --- | --- | --- | --- | --- | --- |
| Age at Surgery,<br>Mean (SD) | 55 (20) | 51 - 59 | 69 (10) | 65 - 72 | 49 (20) | 44 - 53 |
| Female, % (n) | 53% (56) | 43 - 63 | 70% (23) | 51 - 84 | 45% (33) | 34 - 57 |
| AffectedSide,<br>% (n) |  |  |  |  |  |  |
| Axial | 3.8% (4) | 1.2 - 9.9 | 0% (0) |  | 5.5% (4) | 1.8 - 14 |
| Left | 50% (53) | 41 - 59 | 61% (20) | 42 - 77 | 45% (33) | 34 - 57 |
| Right | 46% (49) | 37 - 56 | 39% (13) | 23 - 58 | 49% (36) | 38 - 61 |
| Surgeon, % (n) |  |  |  |  |  |  |
| A | 54% (57) |  | 58% (19) |  | 52% (38) |  |
| B | 41% (43) |  | 42% (14) |  | 40% (29) |  |
| C | 1.9% (2) |  | 0% (0) |  | 2.7% (2) |  |
| D | 3.8% (4) |  | 0% (0) |  | 5.5% (4) |  |
| <sup>1</sup>RevisTHA = RevisionTHA |  |  |  |  |  |  |
| <sup>2</sup>CI = Confidence Interval |  |  |  |  |  |  |

\*One primary presentation of femoroacetabular osteoarthritis with periacetabular bone loss in the setting of hip dysplasia

**Table 2a:** TumourPelvis Cohort (N = 73) frequency of indications for custom printed pelvic implant (%, n).

| Indication | % | n |
| --- | --- | --- |
| Sarcoma | 45 | 33 |
| Other Tumour<br>- Bone giant cell tumour<br>- Tenosynovial giant cell tumour (TGCT\|PVNS)<br>- Neurofibromatosis<br>- Lymphoma<br>- Hemangiomatosis | 22 | 16 |
| Myeloma | 3 | 2 |
| Metastasis to pelvis<br>- Pelvic organs<br>- Renal<br>- Breast<br>- Skin<br>- Lung<br>- Testicular<br>- Prostate | 32 | 23 |
| Radiation-induced necrosis pelvic bone | 18 | 13 |
| Revision of previous tumour reconstruction | 8 | 6 |

\*One case of primary dysplasia not included

**Table 2b:**
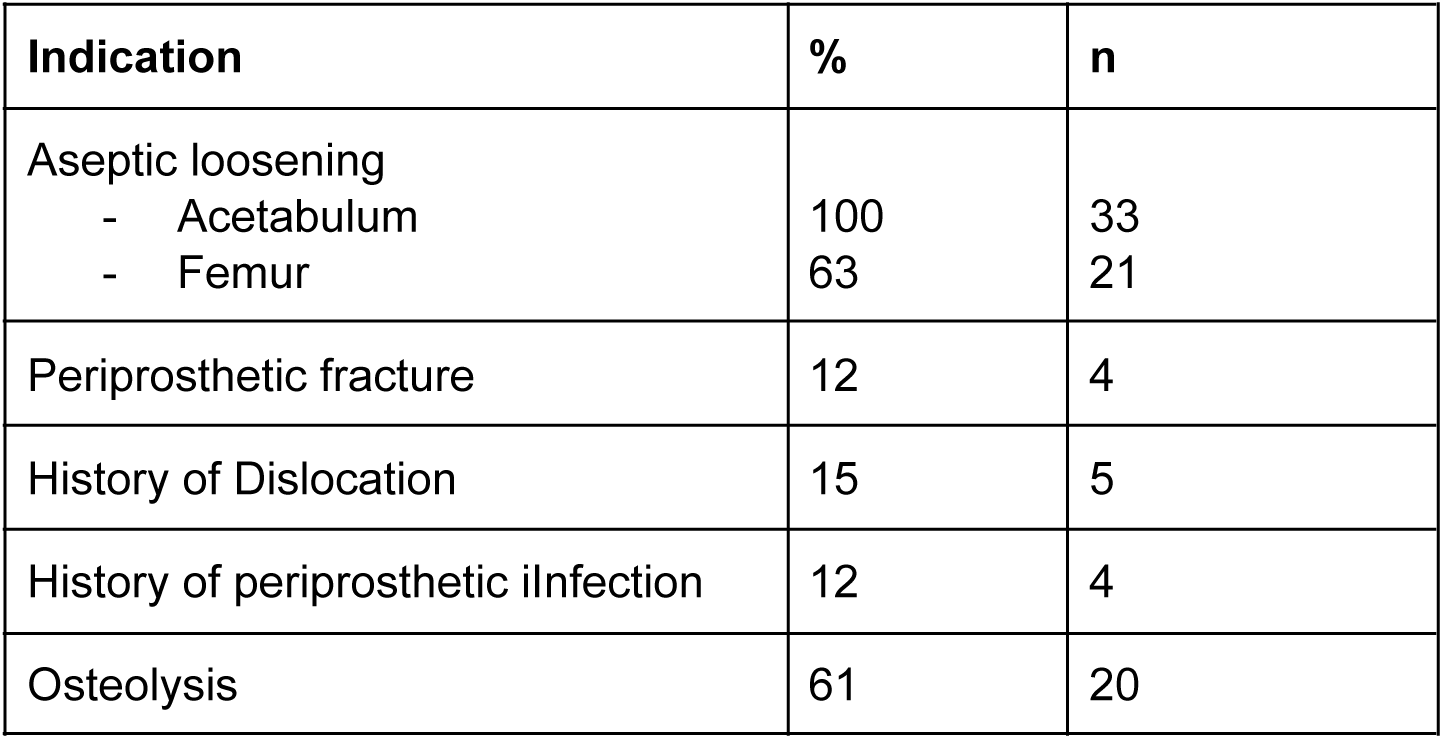
Revision Hip Cohort (N = 33) frequency of indications for custom printed pelvic implant (%, n).

| Indication | % | n |
| --- | --- | --- |
| Aseptic loosening |  |  |
| - Acetabulum | 100 | 33 |
| - Femur | 63 | 21 |
| Periprosthetic fracture | 12 | 4 |
| History of Dislocation | 15 | 5 |
| History of periprosthetic infection | 12 | 4 |
| Osteolysis | 61 | 20 |

### Followup

The overall median follow up of the series was 4.1 (IQR 2.4, 5.6) years, with a range of 0.6 - 10 years.

### Complications and Adverse Events

There were no intraoperative complications, or cases of mortality, tumour or thrombosis in the Reintervention THA cohort (Table 3). Infections were observed (N = 5 (15)%, 95%CI 5.7 - 33%), as well as neurological issues - usually due to screw placement (N = 3 (9.1)%, 95%CI 2.4 - 25%).

**Table 3:**
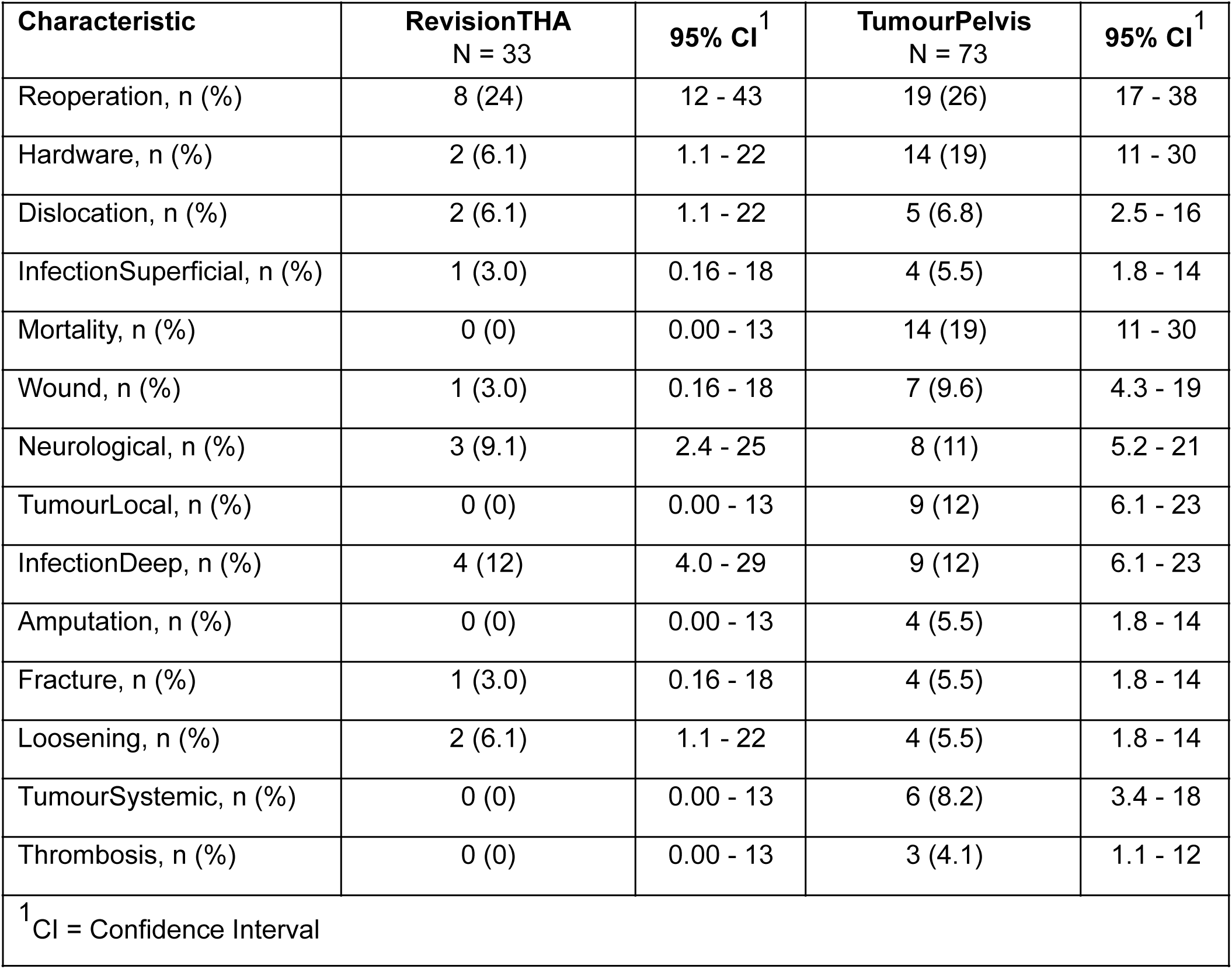
Summary of adverse events after receiving implant of interest (index procedure) by cohort.

There were 3 instances of intraoperative complications (4.1%, 95%CI 1.1 - 12.3) in the TumourPelvis cohort. There was one major intraoperative haemorrhage, one confirmed positive margin and one possible positive margin detected by histopathology. All three cases remained alive with the implant in-situ at the end of the follow up period. The Tumour Pelvic cohort displayed mortality of N = 14 (19)% with relatively wide confidence intervals (95%CI 11 - 30)%, as well as infection (N = 9 (12)%, 95%CI 6.1 - 23%), local tumour recurrence (N = 9 (12)%, 95%CI 6.1 - 23%) and other hardware fixation issues (N = 14 (19)%, 95%CI 11 - 30%) (Table 3).

### Survival Analysis

The analysis demonstrated the change in survival estimates over time for both cohorts. In the RevisionTHA cohort, periprosthetic infection and hip dislocation displayed increased presentation beyond one year follow up (Table 4). Cumulative survival free of adverse events displayed similar patterns for both cohorts for periprosthetic infection (Figure 2) and dislocation (Figure 3). However, the TumourPelvis cohort displayed instances of dislocation within first year of follow up (Table 5), with a relatively linear risk of local tumour recurrence (Figure 4, Table 5).

**Table 4:**
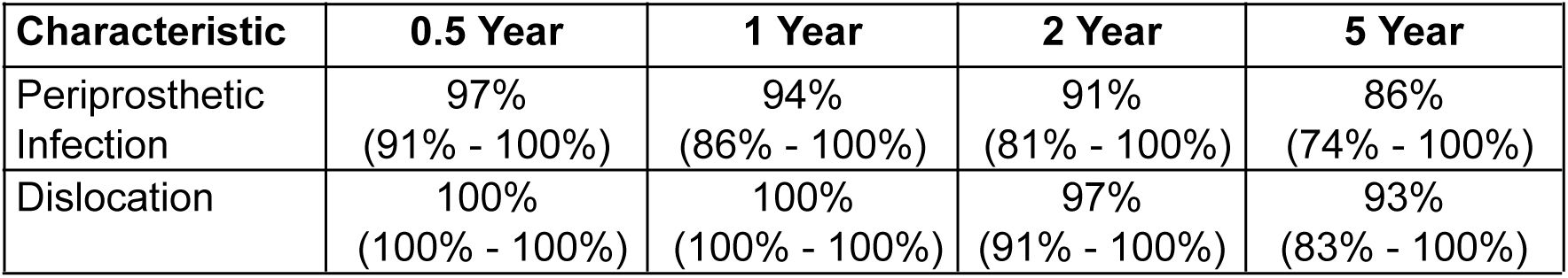
Summary of Kaplan-Meier survival estimates for RevisionTHA cohort.

**Table 5:**
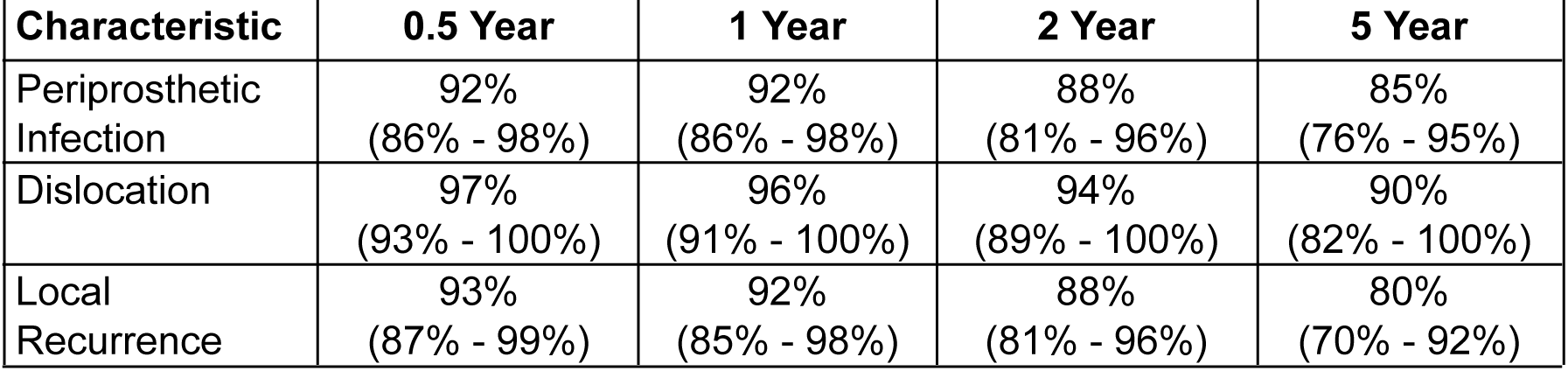
Summary of Kaplan-Meier survival estimates for TumourPelvis cohort.

| Characteristic | 0.5 Year | 1 Year | 2 Year | 5 Year |
| --- | --- | --- | --- | --- |
| Periprosthetic Infection | 92%<br>(86% - 98%) | 92%<br>(86% - 98%) | 88%<br>(81% - 96%) | 85%<br>(76% - 95%) |
| Dislocation | 97%<br>(93% - 100%) | 96%<br>(91% - 100%) | 94%<br>(89% - 100%) | 90%<br>(82% - 100%) |
| Local Recurrence | 93%<br>(87% - 99%) | 92%<br>(85% - 98%) | 88%<br>(81% - 96%) | 80%<br>(70% - 92%) |

**Figure 2:**
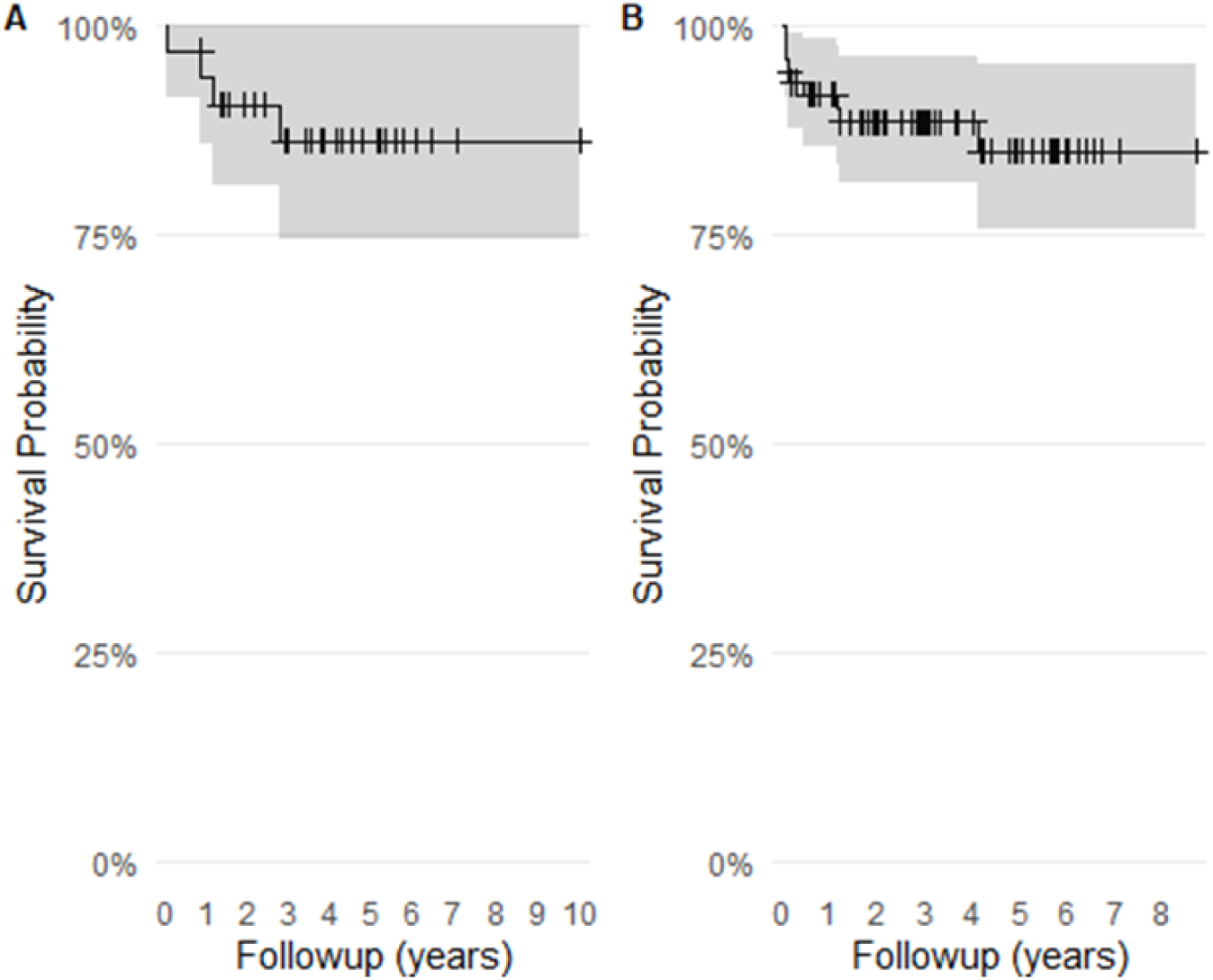
Kaplan-Meier survival curves for Infection in RevisionTHA (A) and TumourPelvis (B) cohorts

**Figure 3:**
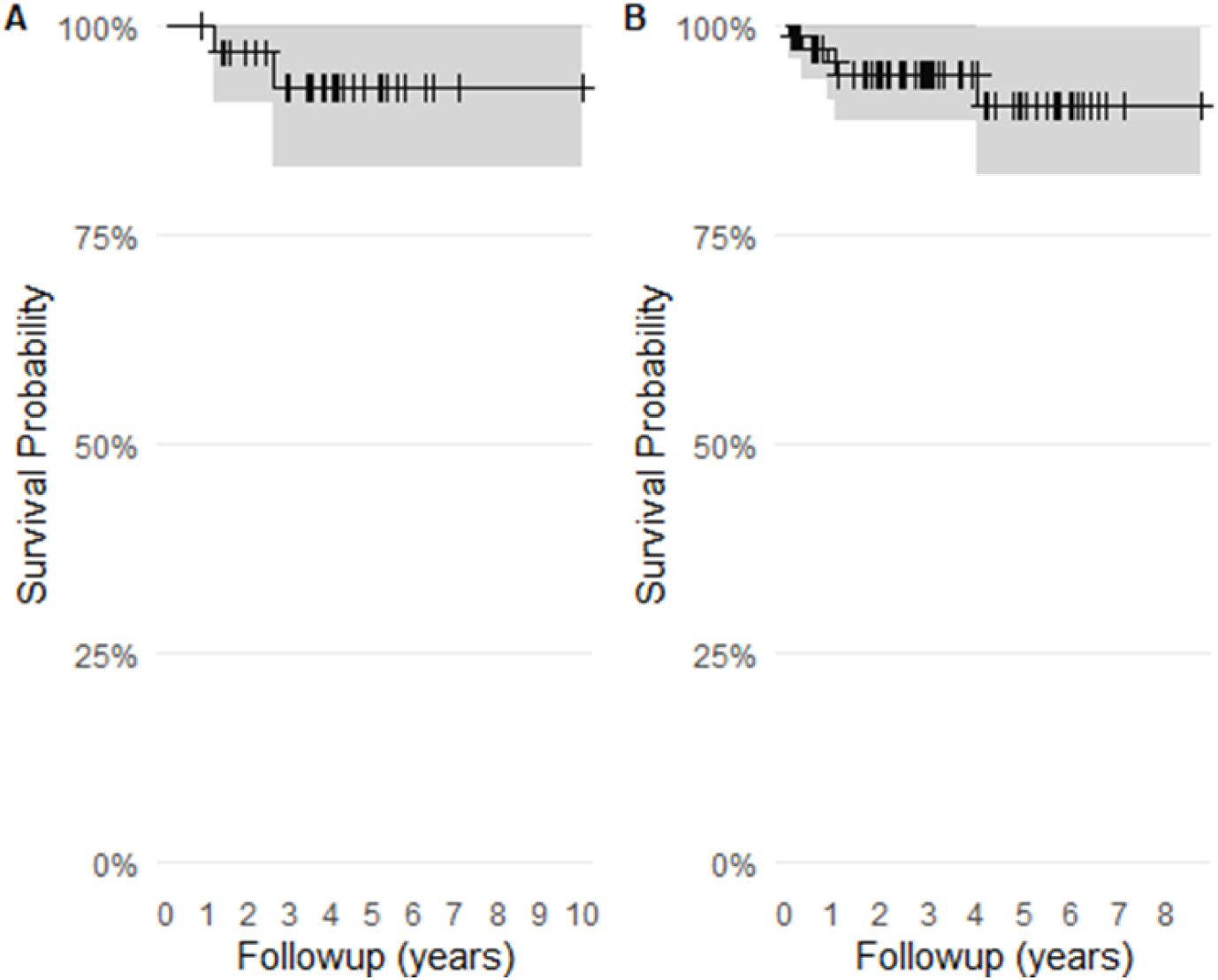
Kaplan-Meier survival curves for Dislocation in RevisionTHA (A) and TumourPelvis (B) cohorts

**Figure 4:**
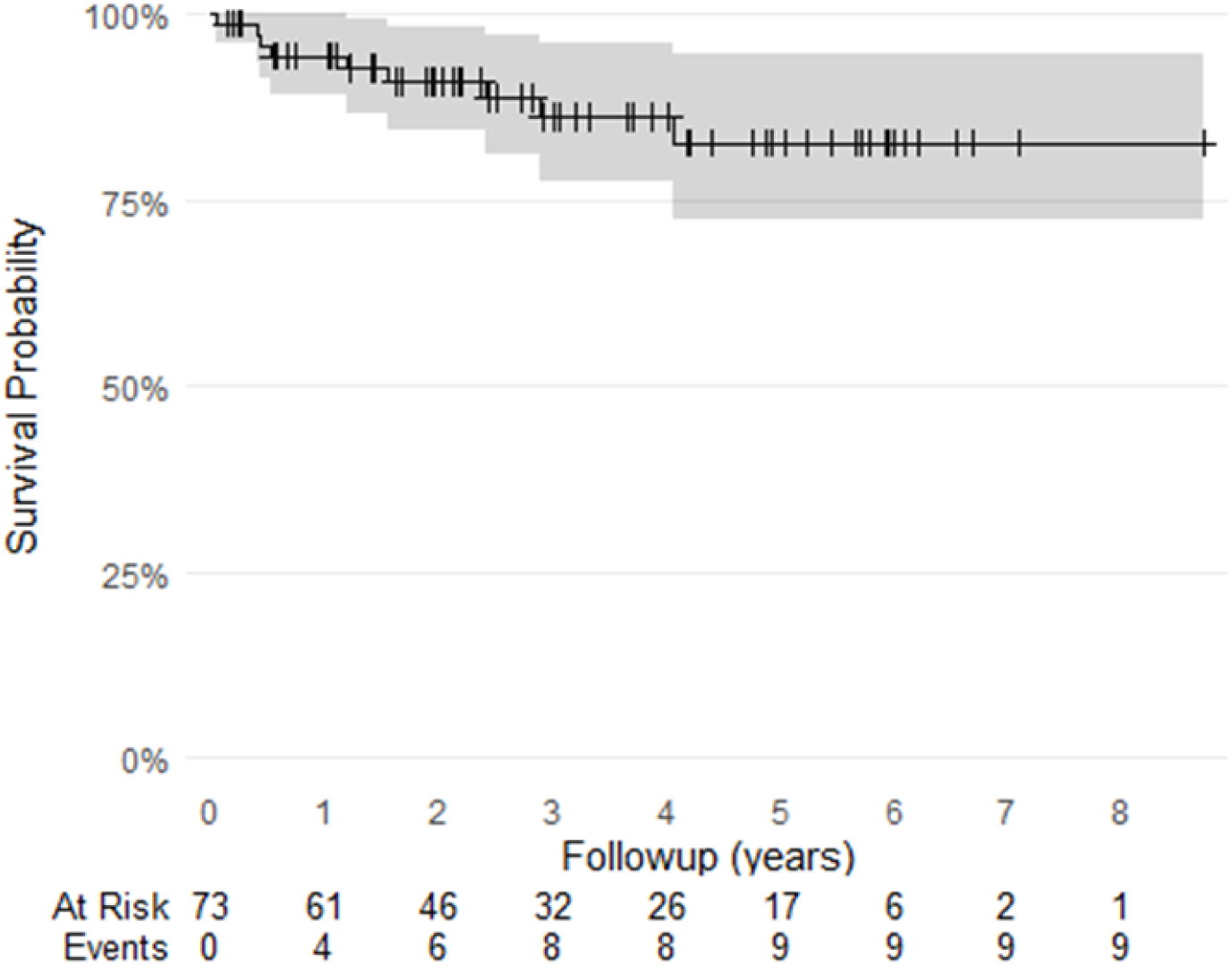
Kaplan-Meier survival curve for Local Tumour Recurrence in TumourPelvis cohort

### Patient survival

No mortality events were observed in the RevisionTHA cohort. Overall, patient survival in the TumourPelvis cohort was 90% at one year, which decreased to 79% by the 5 year follow up (Table 6). The pattern of mortality in this patient cohort was relatively linear up to the three year follow up (Figure 5).

**Table 6:**
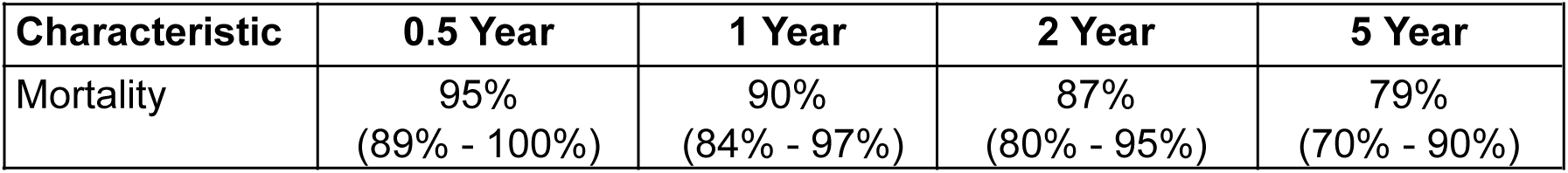
Summary of Kaplan-Meier survival estimates for TumourPelvis cohort.

**Figure 5:**
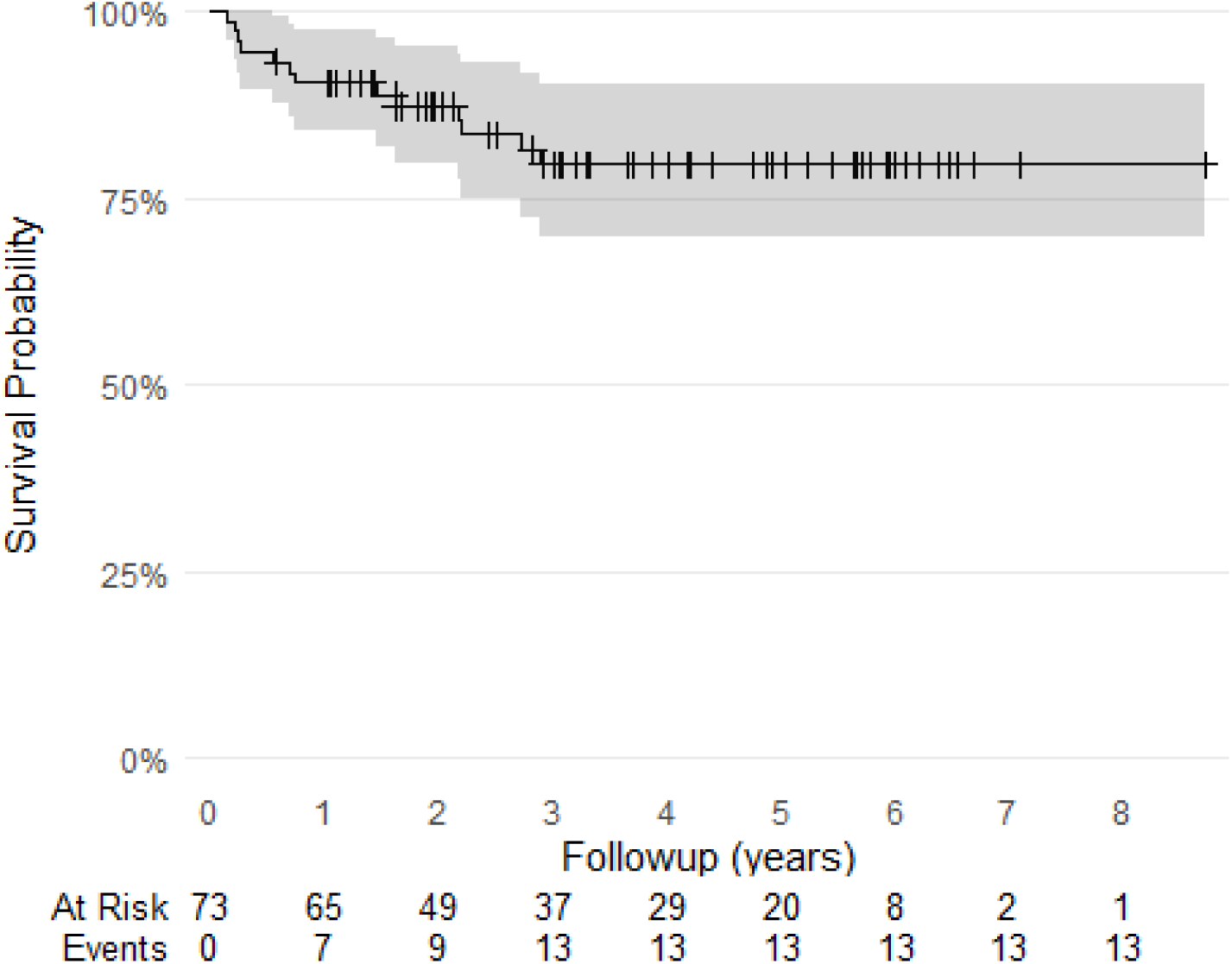
Kaplan-Meier survival curve for Mortality outcome in TumourPelvis cohort

### Implant Retention and Failure

Overall, the implant retention rate for the series was (96.2%, 95%CI 90.1 - 98.8). The pelvic implant was removed in four cases in the TumourPelvis cohort. One component was replaced via two-stage revision for periprosthetic infection. Unfortunately, the patient had metastatic disease progression and subsequently died. One case suffered tumour recurrence and periprosthetic infection and underwent implant removal and hip disarticulation with no further events. Two further cases represented with tumour recurrence and underwent implant removal and hip disarticulation, with one dying. These cases began to represent at one year follow up (Figure 6). There were nine cases considered failed, such that the implant required removal, or any part of the construct required a revision procedure, summarised in Table 7.

**Table 7:**
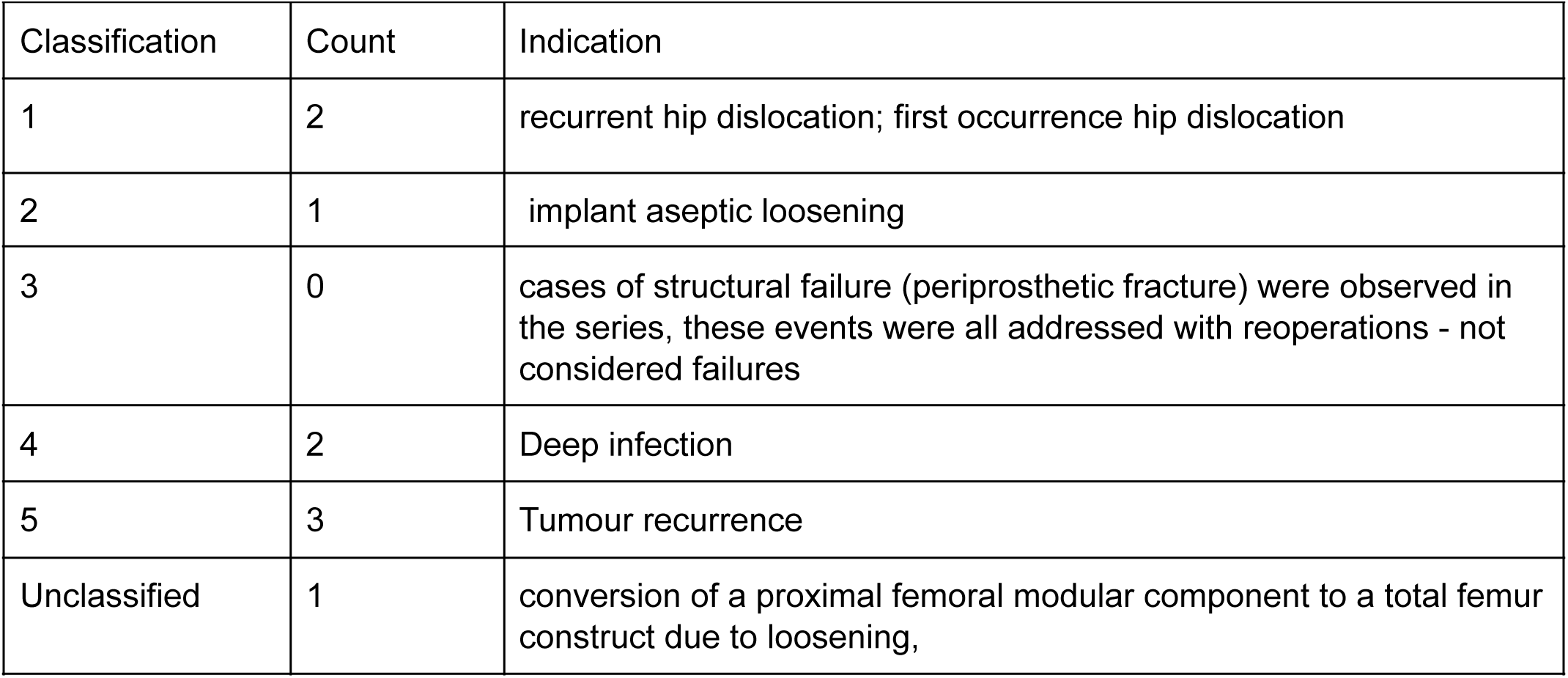
Summary of Henderson criteria for failures.

| Classification | Count | Indication |
| --- | --- | --- |
| 1 | 2 | recurrent hip dislocation; first occurrence hip dislocation |
| 2 | 1 | implant aseptic loosening |
| 3 | 0 | cases of structural failure (periprosthetic fracture) were observed in the series, these events were all addressed with reoperations - not considered failures |
| 4 | 2 | Deep infection |
| 5 | 3 | Tumour recurrence |
| Unclassified | 1 | conversion of a proximal femoral modular component to a total femur construct due to loosening, |

**Figure 6:**
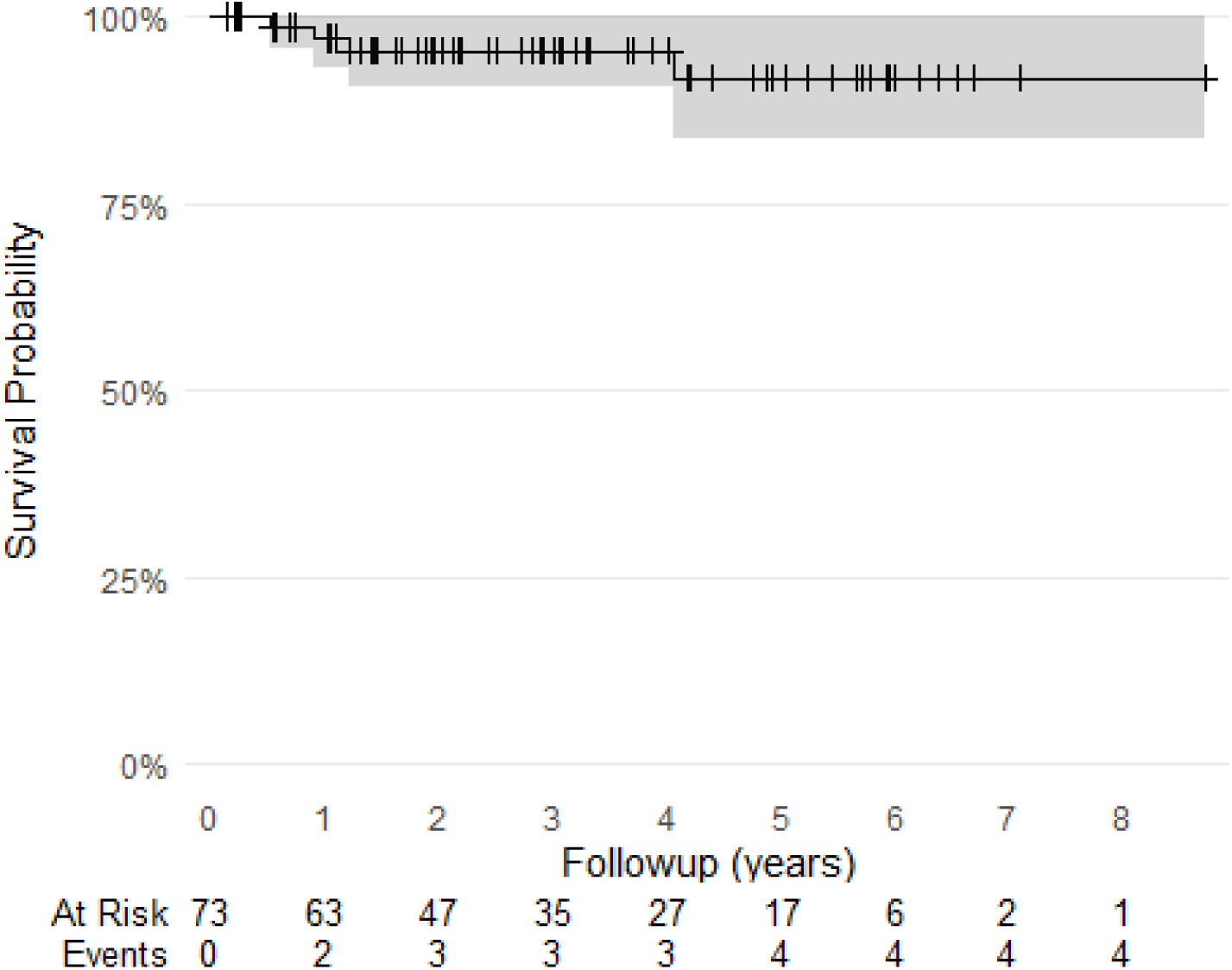
Kaplan-Meier survival curve for Amputation | Implant Removal in the TumourPelvis cohort

The multistate model illustrated the complex nature of adverse events in this population, particularly for the TumourPelvis cohort. Cumulative incidence of adverse events within the context of a multistate model identified the increasing burden of reoperations required for TumourPelvis cases following index implantation up to the one year follow up (Figure 7), which diminished over time. An oscillating pattern of cumulative incidence was also observed (Figure 7 - bottom), with patients remaining in an adverse event state for extended periods for some complication types (e.g. periprosthetic infection), while for other types (e.g. dislocation) they transitioned relatively quickly.

**Figure 7:**
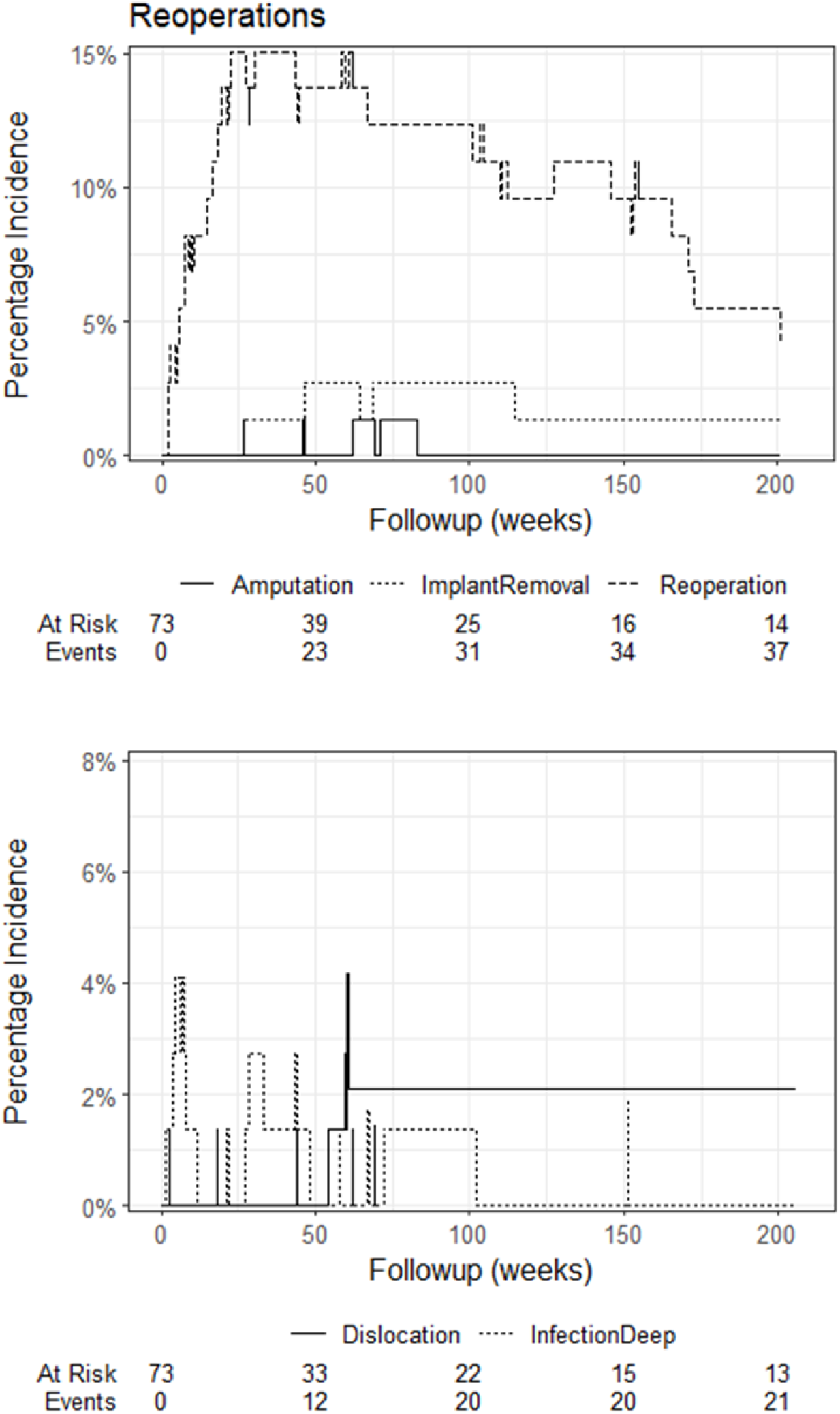
Incidence of reoperations (top) and infection|dislocation (bottom) over time for Tumour cases.

## Discussion

Pelvic reconstruction using custom-designed, three-dimensional printed triflange implants (3DPI) for severe bone defects in the context of failed total hip replacement or tumour resection remains an emerging area. The present understanding of the clinical outcomes of these types of implants remains constrained by small-sample series for both revision THA (Broekhuis et al., 2023; De Martino et al., 2019) and pelvic tumour resection (Hu et al., 2023). The aim of the present study was to conduct the first comprehensive review of the cases performed in our unit for both indications and report the incidence and timing of adverse events. This study builds on previous work from our group that reported the clinical outcomes of a subsample of tumour resections (Broekhuis et al., 2022) and the anaesthetic-related challenges in this mixed population (Chua et al., 2019).

Appropriate patient selection and the transparent assessment of risk in cases appropriate for 3DPI remains an ongoing challenge for the treating surgeon and the patient. The results of the present study provide additional information to the consideration of risk versus benefit in this population with regard to the use of 3DPI, particularly in the setting of tumour resection. A key finding was 10% mortality (3 - 16%) by 12 months follow-up in the tumour cohort. Although a guiding principle in the recommendation for 3DPI has been an expected favourable patient prognosis within the 12 months from the intended date of surgery, it appears this may not be realistic for approximately 1 in 10 patients treated historically within our unit. The very few efforts that have been directed to develop prognostication models for sarcoma and limb-salvage mortality and disease-free survival (Zheng et al., 2021), require further external validation and are likely underpowered for precise estimates (Riley et al., 2019). The present cohort likely remains below sufficient sample size for external validation (Riley et al., 2022). Only through multicentre collaboration and careful prospective data collection will sufficient data be collated to enable localised prediction for future pelvic resection patients.

In contrast, the issue of patient selection for revision THA is somewhat different, with no mortality events observed in the present study. Therefore, the considerations naturally shift to one of implant longevity and functional restoration for the patient. In this, the present results are less definitive, with multiple adverse events observed in some patients, but of a less complex nature than the tumour resection cohort and the overall implant survival was acceptable. Past attempts to prognosticate 3DPI implant longevity in this cohort have been limited in scope and constrained with respect to the available sample size (Barlow et al., 2016). Overall, in the patients that would most benefit from prognostic tools to enable counselling of expectations and make informed decisions with regard to risk and benefit, heterogeneity of indications combined with relatively low case volume continue to pose significant challenges to the availability of tools fit for routine clinical implementation.

One of the most devastating complications of 3DPI is infection in both pelvic tumours (Angelini et al., 2013) and revision THA (De Martino et al., 2019), which is difficult to manage and can result in the need for hindquarter amputation with or without implant removal in certain circumstances. Direct comparisons to the contemporary literature should be made with caution due to i) lack of clarity with respect to diagnostic criteria and definitions across the related literature, ii) ignorance of time-to-event (including in systematic reviews) and case censoring in previous studies, ii) ignorance of competing or multiple concurrent events when calculating incidence in previous studies and iii) insufficient sample size for comparison in the present series (see limitations). In this study we observed a rate of periprosthetic infection of 8% with a confidence interval of 2 -18% in tumour cases by 12 months follow-up. The present series illustrates the evolution of care in the pelvic tumour population, with an earlier series (Angelini et al., 2013) reporting 17% infection incidence at one year followup, and of those, nearly half (46%) required implant removal. The present series demonstrated a much higher implant retention rate in infected cases. Our results also compare favourably to a contemporary series from Japan which reported 32% infection incidence in 65 cases with custom triflange for tumour resection (Fujiwara, Medellin Rincon, et al., 2021), although the time-to-event data was not reported. The present results are also comparable to a nationwide survey of pelvic resection cases (Mavčič et al., 2020), that reported a cumulative incidence of infection of 12.5% (no follow up threshold provided). In contrast, a recent series of pelvic tumour reconstructions (N = 96) reported a rate of 4.2% (Hu et al., 2024), although on closer inspection, the difference to the present results may be attributed to stricter definition of an infection event in the Hu paper. The revision THA cohort in this study displayed an infection incidence of 6% (95%CI 0 -14%) by 12 months follow-up. This rate is somewhat comparable to (Broekhuis et al., 2023) who reported an incidence of 3.3 - 6.5% from a systematic review of 1218 procedures in 33 studies. An earlier systematic review of custom triflange acetabular components in revision THA reported an overall infection incidence of 6.5% (De Martino et al., 2019) from 579 procedures (17 studies). The broad equivalence between the literature and the present study should be interpreted cautiously for the reasons stated above.

Although the infection rate by 12 months follow up appears comparable to the literature, the analysis reveals an increasing incidence over time. In both indication types, cumulative incidence by 5 year follow-up approximately doubles from 12 month follow-up. 3DPI cases remain vulnerable to periprosthetic infection and haematological seeding remains a considerable threat to implant viability into the medium-term. These infections may be predominantly polymicrobial and caused by gram-negative microorganisms associated with intestinal flora (Sanders et al., 2019). Further, these results prompt three further courses of enquiry. The first is that understanding the time course of adverse events, as shown by the survival analyses presented for other categories, is an important aspect of improving future clinical practice in terms of informed patient counselling, as well as establishing effective patient follow up timetables and monitoring methods. The second is to inspire improved standards of analysis of these patient populations and evolve the discourse around adverse events after implantation of the 3DPI.

Whilst there is increasing recognition of competing events (Fujiwara, Medellin Rincon, et al., 2021) and need for multistate models when evaluating outcomes (Schuster et al., 2020), these approaches remain uncommon in contemporary literature, where even time-to-event as a concept remains largely ignored (Broekhuis et al., 2023; Hu et al., 2024). The third avenue is to further refine perioperative and postoperative management to continue driving down infection incidence, in both the acute and chronic categories. Previous work by our group has demonstrated the complex nature of pelvic tumour resection patients (Chua et al., 2019), with long average procedure duration and large amount of blood loss compared to other elective joint procedures such as knee and hip arthroplasty. Known risk factors for post-surgical infection include intraoperative blood loss (Aeschbacher et al., 2021), transfusion (Kim et al., 2017), and perioperative anaemia (Zhang et al., 2024), as well other patient characteristics such as pre-existing comorbidities. Prophylactic antibiotics protocols administered acutely may be one element of continued research, increased attention to preoperative patient optimisation for surgery and medical management over the medium-term, with attention to intestinal microbiology (Sanders et al., 2019) may be just as effective in further reducing infection rates.

The present review of adverse events revealed neurological issues (palsy, sciatic irritation) in approximately 10% of cases, as well as hardware issues pertaining to screw fixation in 6% of revision cases and 19% in pelvic tumours. In many cases, these issues could be attributed to misplacement of screws relative to the preoperative plan. Work to date on assessment of implant accuracy is yet to investigate positioning deviations as they relate to screw trajectories (Broekhuis et al., 2024). Although neural structures are accounted for in planning for screw placement, subtle malposition of the 3DPI can be particularly apparent in pelvic tumour cases where resection of the affected segment often releases hoop tension within the girdle and can lead to intersegmental shift relative to the preoperative plan. This can contribute to misplacement of fixation screws with respect to the target segment relative to the implant as well as its relationship with key nerve structures such as the sacral nerve roots. Deviations in the screw trajectory can lead to nerve irritation, as well as screw loosening or breakage. Future efforts could be directed towards using guidance systems to maintain awareness of segments during the procedure. Computer navigation has been advocated in the past for this purpose (Christ et al., 2021; Dang et al., 2022; Fujiwara, Kaneuchi, et al., 2021) and has shown benefit with respect to resection accuracy, however there remain concerns over extending surgical time for already length procedures, as well as the potential for additional complications associated with intracortical trackers. The introduction of augmented and mixed reality systems may alleviate some of the issues with navigation in pelvic tumour resection, but development for broad clinical adoption remains in its early stages (Sun et al., 2023).

The present analysis contributes additional knowledge to the use of 3DPI in general in the treatment of severe pelvic bone defects associated with revision THA and pelvic tumour resection. The results also verify the clinical utility of the preoperative planning and device manufacturing utilised by our unit specifically. Clinicians and researchers can apply the information to better guide practice with respect to preoperative counselling and postoperative monitoring and management, However, the results should be interpreted within the context of the limitations of the study. Firstly, the patient cohorts reviewed separately constrain the precision with which time-to-event data can be reported, with particular reference to the width of the confidence intervals around survival estimates for various adverse event categories. The available sample size remains insufficient to perform definitive comparisons to benchmarks within the literature or conduct more detailed inference within the sample. Secondly, the broad case heterogeneity within indication cohorts further impacts on the precision within the available samples. Thirdly, As with any retrospective review, biases with respect to the information available and the ability to accurately classify cases into the correct indication group cannot be ruled out. Information bias is present in the evolution of information management systems over time within participating practice as well as ancillary systems (e.g. laboratory and imaging). Classification bias is partly related to this, with some cases much clearer to classify than based on the available information, however in some tumour cases with a past history of resection and reconstruction with or without arthroplasty, the ability to classify into revision THA or pelvic tumour is challenging. Future work utilising multicentre, prospective and organised data collection combined with data simulation is required to enable more definitive interpretation to achieve sufficient analytical samples with precision and to mitigate biases associated with retrospective observational reviews.

## Conclusion

3DPIs are a safe and viable option for complex reconstruction of the pelvis across a range of oncological and non-oncological indications. The initial results of the present study provide important information to aid in counselling patients about such procedures and allocating healthcare resources for ongoing care. Further work is required to document functional and biomechanical outcomes in these patient populations.

## Data Availability

All data produced in the present study are available upon reasonable request to the authors

## Acknowledgements

Natalie Bennell (Ossis Ltd) for her assistance with the vendor case list for crossmatching.

Anthony Maher for his assistance with data retrieval.

EBM Analytics for maintenance of the COMPRESSOR registry system.

Patients and staff of the contributing practices.

