## Supplementary 2 for "High retention rates of custom 3D printed titanium implants in complex pelvic reconstruction, a report on 106 consecutive cases over 10 years"

### Three-dimensional printed pelvic implants for oncologic and non-oncologic indications: Mortality, procedure survival and adverse events in a consecutive series

Corey Scholes, Chief Science Officer EBM Analytics

2024-08-30

10.1 STROBE [13.1] Treatment selection ... 17
11. STROBE [14] Patient and record characteristics ... 17
11.1 STROBE [14.1] Pathology characteristics ... 18
11.2 STROBE [14.2] Management summary ... 20
11.3 STROBE [14.3] Follow up ... 20
12. STROBE [15] Outcomes ... 22
13. STROBE [16] Main results ... 22
13.1 Kaplan-Meier survival curves ... 23
13.2 Multi-state survival model ... 26
13.3 Henderson Failure Criteria ... 33
14. STROBE [17] Sensitivity analyses ... 33
15. Ethical Considerations: ... 33
15.1 Informed Consent: ... 33
15.2 Ethical Approval: ... 33
15.3 Privacy and Confidentiality: ... 34
16. Conflicts of Interest: ... 34
16.1 STROBE [22] Funding: ... 34
16.2 Disclosure: ... 34
16.3 Potential for Biases: ... 34
17. Analysis References ... 34

### 1. Introduction

This analysis links to the manuscript of the 3DPI project.

#### 1.1 Preparation

Load up required packages in advance; check if packages are installed, if not, install them. Citations applied to each library at first use in the text.

#### 1.2 Aim

To describe the clinical outcomes, in patients presenting for surgical review of pathologies leading to bone defects through bone resorption or surgical resection of the pelvis appropriate for treatment with three-dimensional, customised printed pelvic implants.

#### 1.3 Hypothesis

The following analysis is descriptive only. No hypotheses have been constructed for this case series.

### 2. Analysis Methods

#### 2.1 Reporting

The study was reported according to the STROBE guidelines (Vandenbroucke et al. 2007) and companion checklist. The analysis was conducted in RStudio IDE (RStudio v2024.04.1+748 “Chocolate Cosmos” Release ) and Quarto (v1.5.17) using R (vR version 4.4.0 (2024-04-24 ucrt)) and associated packages to perform the following;

- Data import and preparation
- Sample selection
- Describe missingness
- Data manipulation and analysis of;
  - Patient characteristics
  - Pathology characteristics
  - Management details
  - Procedure survival, adverse events and complications
- Survival analysis - single-event
- Survival analysis - multistate
- Tabulation and presentation

#### 2.2 STROBE [4] - Study Design

Subgroup analysis of a clinical registry embedded into private practice (COMPRESSOR). Observational, cohort design. Cases added retrospectively into registry were aligned with schema based on electronic chart review.

#### 2.3 Data Import and Preparation

Retrieve and format data from live tables and registry snapshot. Using *openxlsx* (v1.8) (Barbone and Garbuszus 2024) to retrieve static snapshot files and *googlesheets4* (v{r}``utils::packageVersion``(pkg = "googlesheets4")) (Bryan 2023) to retrieve live database tables. Text and code output are integrated using the *epoxy* package (v1.0.0) (Aden-Buie 2023).

Combine dataframes into one to conduct analysis using *dplyr* (v1.1.4) (Wickham et al. 2023). PatientID was extracted from TreatmentID using *stringr* (v1.5.1) (Wickham 2023) applied over the dataframe using *purrr* (v1.0.2) (Wickham and Henry 2023) and a combination identifier was created by combining PatientID and AffectedSide to traverse across treatment records.

##### 2.4 STROBE [5] - Setting

The COMPRESSOR registry is based in a mixed (public | private) practice for ortho-oncology and complex upper and lower limb arthroplasty located in Sydney, Australia. It contains a mix of retrospectively identified records retrieved through chart review, as well as prospective recruitment of patients on initial presentation to the clinic for surgical review.

The registry has 1312 treatment records with the first patient added 27 November 2020 and the final treatment record created 02 August 2024.The registry snapshot was extracted on 01 August 2023.

#### 3. STROBE [6] Participants

##### 3.1 STROBE [6.1] Sample selection

Identify cases receiving the implant of interest. Cases were identified by the manufacturer tag in the AnalysisLabel field of the TreatmentTable of the registry database.

Inclusion criteria for the AnalysisLabel field;

- Case involves implant of interest
    - offered (and declined by the patient)
    - planned (implant manufactured, but not implanted)
    - or implanted
- Case of interest in this analysis is the index procedure within the registry (first use of implant)
- Patient has not withdrawn consent for inclusion of data in the registry
- Treatment STROBE is eligible for surgery (whether surgery occurred or not) with the implant of interest

Data manipulation (add columns and filter tables based on column values) was performed with *tidyverse* (v2.0.0) (Wickham et al. 2019), and *dplyr* (v1.1.4) (Wickham et al. 2023) and conversion to display format using *gt* (v0.11.0)(Iannone et al. 2024).

Use AnalysisLabel to identify treatment records receiving (or offered) implant of interest.

Write to CLC rawData for processing

Cycle through mastersheet table and complication processing

Retrieve all treatments for index *patients* and return a table

##### 3.2 STROBE [6.2] Algorithm validation

STROBE selection code was cross-checked by manual STROBE checking within the registry snapshot for a subset (N = 10) of cases.

##### 3.3 STROBE [6.3] Data linkage

Where additional review of adverse event data was conducted with the attached hospital records, patients were identified by name, date of birth and date of surgery. Manual review was performed by a resident familiar with the system and the case mix of the participating consultant surgeons.

#### 4. STROBE [7] Variables

Key variables defined as part of this analysis are summarised in Table 1 below.

Table 1: Summary of variables presented in analysis of 3-dimensional printed pelvic implants

| Category | Variable | Definition | Comments |
|---|---|---|---|
| Inclusion-Exclusion | Label | AnalysisLabel of registry STROBE contains "Ossis" | |
| | IndexProcedure | First instance of use for implant of interest | |
| Patient Characteristics | Age | Biological age on the date of IndexProcedure | |
| | Sex | At the time of surgery; sex transition was not captured in the dataset | |
| | AffectedSide | Hemisphere of the pelvis in which the centre of mass of the tumour is located | Axial was added as a value for cases where the tumour is located predominantly in the sacrum |
| Pathology | DiagnosisRaw | Raw text field where electronic records are distilled to a description of the relevant diagnostic | |

| Category | Variable | Definition | Comments |
| --- | --- | --- | --- |
| | | information for a given case | |
| Management Details | Surgeon | The primary consultant surgeon conducting the procedure | |
| Adverse Events | Mortality | Description of death included in medical STROBE | |
| | ImplantRemoval | Instance whereby the IndexProcedure component is ultimately removed from the patient | equivalent to standard definition for joint arthroplasty |
| | Key events | Infection - documented cases of infection (positive cultures) OR *suspected* whereby indication for reoperation is predicated on an assumption of infection<br><br>Dislocation - document instance of separation of femoral head from acetabulum cup<br><br>Loosening - documented migration of the implant of interest or related components<br><br>Amputation - documented disarticulation of the femoral head and removal of the leg from the hip level | Amputations did not lead to component removal |
| | Tumour Recurrence | Local - presence of new tumour growth within surgical field<br><br>Systemic - recurrence or flare of non-regional specific tumour (e.g. myeloma) or to area outside of surgical field | |

| Category | Variable | Definition | Comments |
|---|---|---|---|
| | | (metastasis) originating from surgical site or elsewhere. | |
| | Recurrent events | Wound issues - dehiscence or other breakdown of the surgical wound<br><br>Fracture - Fracture of the pelvis<br><br>Thrombosis - Any documented blood clotting event<br><br>Neurological - Documented symptoms reported after surgery associated with control or sensation of the lower limbs or organs<br><br>Hardware - loss of fixation or other issues with component screws | Fixation of ASIS fragment |
| Survival Analysis | Duration | Difference (in weeks) between the date of surgery and the date of event occurrence | Separated into Start and Stop (date of next sequential event for patient STROBE) |
| | EventCategory | Label for each adverse event entry in the table corresponding to a category Generated from text pre-processing and some manipulation of key terms (addition) to match regular expressions used for presentation | Some free text entries of complication nature required amendment to fit regex |

#### 5. STROBE [8] Data sources

Data was retrieved from chart review and;

- Entered into the database through the registry interface

- Complications and adverse events captured into an online form (QuestionPro, USA) and linked using STROBE identifier codes.

#### 5.1 Adverse Events

Complication entries were written to an external file for co-author review.

The free-text describing the nature of the complication or adverse event was pre-processed using *tidytext* (v0.4.2)(Silge and Robinson 2016) to split into word tokens and remove stop words.

Terms with less than four characters were extracted and reproduced in an external file for manual spelling of abbreviations. Terms with digits (e.g. L5) were removed.

The abbreviated terms with expanded definitions were read back into the workspace for replacement in the complication descriptions.

The terms were replaced using *stringr* functions and added to the dataframe containing complication data.

A figure displaying term frequency was generated using *ggplot2* (v3.5.1)(Wickham 2016) and formatted for reporting using *knitr* (v1.48) (Xie 2024).

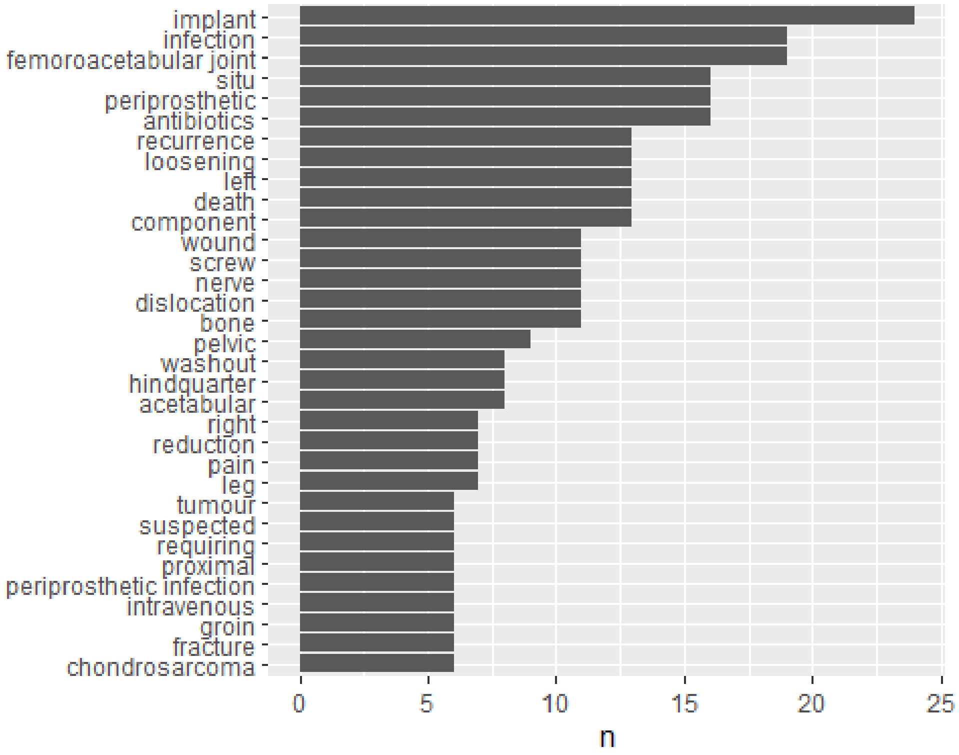

Figure 1: Complication terms by frequency

A wordcloud was generated using *wordcloud* (v2.6) (Fellows 2018) to express the most common terms in the complication description free text field.

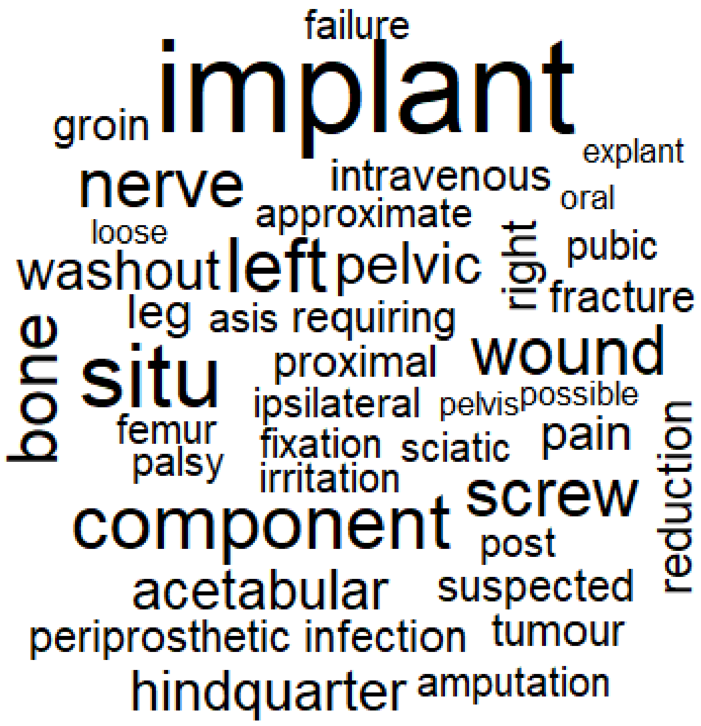

```
NULL
```

Figure 2: Word cloud of top 50 terms in the complication description data entry

#### 6. STROBE [9] Bias

For a discussion of biases in the context of the clinical registries in general, refer to (Scholes et al. 2023). Specific to this analysis, the following considerations are noted;

Table 2: Summary of potential biases in the described analysis

| Bias | Definition | Source | Mitigation |
|---|---|---|---|
| Information | Data from different sources possess varying levels of quality or detail | (Hammer, Prel, and Blettner 2009; Tripepi et al. 2008) | Can not be fully mitigated; Practices used different systems to store and organise information |
| Misclassification | Treatment STROBE labelled into incorrect cohort.<br><br>Prior treatments of tumour beyond 10 year follow up (discharge) classified | (Benchimol et al. 2015; Tripepi et al. 2008) | Clinical notes reviewed by experienced reviewers and defined within the group a-priori |

| Bias | Definition | Source | Mitigation |
|---|---|---|---|
| | as ReinterventionTHA | | |
| Selection | Treatments are selected based on post-treatment criteria | (Nguyen et al. 2021) | Unable to be mitigated fully - records are identified by presence of implant of interest |
| Censoring - 1 | The first-occurrence of adverse events was specified in the multistate model. In cases where a subsequent event was observed, an event that stays with the patient for life (e.g. infection) may be truncated | | Cannot be fully mitigated - the structure of the survival model requires that subsequent events are organised in sequence and cannot be overlapping |
| Censoring - 2 | Competing events are censored from survival dataset which bias estimates of incidence upwards | (Coemans et al. 2022) | Utilise multi-state survival model (recurring and competing events) |
| Pseudoreplication | Analyse data while ignoring dependency between observations. Inadequate model specification. | (Davies and Gray 2015; Lazic 2010) | Cluster for patient in survival analysis (multistate model). |
| Consent | Patients that tend to opt-in to clinical research can deviate from the population of interest | (Man et al. 2023) | Waiver of consent utilised to access all eligible records |

#### 7. STROBE [10] Sample size

Sample size was derived based on the available records from the Registry at the time of analysis.

#### 8. STROBE [11] Quantitative variables

A number of variables were generated to conduct the survival analysis (see section STROBE 12 below).

Variables from the treatment records of the index procedures were merged with the complication dataframe to ensure validation of the dataset manipulation and to calculate key quantitative variables for survival analysis.

records associated with treatments prior to the index procedure were flagged for exclusion. Regular expressions were used in the *stringr* package to create new columns flagging the presence or absence of the adverse events (see Table 1).

Add reoperations and endpoints to table

The dataset was reshaped to a long format and the indicator columns for each adverse event type were combined into one column within the dataframe (*Category*).

The date of surgery for the index procedure was linked to each complication entry and subsequent quantitative variables such as the durations between;

- date of surgery (index procedure) and date of occurrence
- date of occurrence and date of reoperation
- date of surgery (index procedure) and date of reoperation

Whether the event was intraoperative or presented postoperatively was also assessed using the size of the duration between date of surgery and date of occurrence.

records with no complication recorded, as well as the final period of right-censor for each STROBE that did not undergo removal of the pelvic implant and remained alive at the end of the chart review period (censored) were generated and added to the complication table to enable reorganisation into a format appropriate for the analysis selected.

Censored treatment records (with no complication recorded at all) were combined with records that were censored after one or more complication events to form the *Censored* component of the adverse events dataset.

The censored data records were integrated into the dataset, with the resultant new frame reorganised into a format appropriate for a multi-state model (see STROBE 12.5) of procedure (and patient) survival after implantation with a custom-printed pelvic component, as described in the *survival* package (v3.7.0 ) (Therneau 2024).

A *duration* variable was calculated to arrange the dataframe rows within each PatientID in descending order of occurrence to establish the transition patterns from one health state to the next. The start and stop times for certain events (mortality, amputation) were offset by one *week* to remove ties for recurrent events or different event types occurring on the same date for the same patient. The presence of each adverse event type was restricted to the first occurrence of each Category within a patient subsequent to an index procedure per date of occurrence.

Pre-process for assessment of adverse events survival

```
# A tibble: 0 × 2
# ℹ 2 variables: CombID <chr>, duplicate_dates <lgl>
```

### 9. STROBE [12] Statistical methods

A number of analytical techniques were employed to i) clean the data inputs as well as ii) evaluate missingness in the dataset and iii) complete the descriptive analysis of;

- Patient characteristics
- Pathology details
- Patient, implant and adverse event time to event

#### 9.1 STROBE [12.1] Access to population

The registry system represents all records captured for a surgical group within Sydney, Australia using the implant of interest from its market introduction to present day. All reviewed charts from the operating surgeons practice records (electronic medical record, paper files where required) were entered manually into database and the present analysis draws data from the live database tables directly.

#### 9.2 record [12.2] Data cleaning methods

Diagnosis and complication description free text fields were pre-processed using *tidytext* to remove relational terms (stopwords) and expand abbreviations to improve clarity.

Dates of events (preceding and subsequent surgical records; adverse events including mortality) relative to index surgery date were assessed using coded checks to flag anomalies and were resolved by further manual review to resolve inconsistencies or discrepancies with the chart review input data stored in the registry database.

The dataset used as input for the survival analysis of adverse outcomes was assessed using the *survival* package (v3.7.0) (Therneau 2024), with visual assessment of the transitions table to ensure procedure endstates (mortality, implant removal) do not have subsequent states and that the numbers of events and unique identifiers matched the numbers in the dataframe.

```
Call:
survival::survcheck(formula = Surv(DurationStart, DurationStop, 
    Category) ~ 1, data = ComplicMaster, id = CombID, timefix = TRUE)

Unique identifiers       Observations        Transitions 
               106                301                272 
```

```
Transitions table:
                      to
from                    Wound Hardware Fracture Neurological Dislocation
  (s0)                      5       10        3            7           5
  Wound                     0        0        0            0           1
  Hardware                  0        1        0            3           0
  Fracture                  0        1        0            0           0
  Neurological              0        0        0            2           0
  Dislocation               0        0        0            0           2
  Loosening                 0        0        0            0           0
  Thrombosis                0        0        0            0           0
  TumourSystemic            0        0        0            0           0
  TumourLocal               0        0        0            0           0
  InfectionSyst             0        0        0            0           0
  InfectionSuperficial      0        1        0            0           0
  InfectionDeep             1        1        1            1           0
  Reoperation               3        5        1            0           4
  Amputation                0        0        0            0           0
  ImplantRemoval            0        0        0            0           0
  Mortality                 0        0        0            0           0
  Censored                  0        0        0            0           0
                      to
from                    Loosening Thrombosis TumourSystemic TumourLocal
  (s0)                          3          1              4           5
  Wound                         0          0              0           0
  Hardware                      0          0              0           2
  Fracture                      0          0              0           0
  Neurological                  1          0              1           0
  Dislocation                   0          0              0           0
  Loosening                     0          0              0           0
  Thrombosis                    0          0              1           0
  TumourSystemic                1          0              0           0
  TumourLocal                   0          1              0           0
  InfectionSyst                 0          0              0           0
  InfectionSuperficial          0          0              0           0
  InfectionDeep                 0          0              0           0
  Reoperation                   1          0              1           2
  Amputation                    0          0              0           1
  ImplantRemoval                0          1              0           1
  Mortality                     0          0              0           0
  Censored                      0          0              0           0
                      to
from                    InfectionSyst InfectionSuperficial InfectionDeep
  (s0)                              1                    4             6
  Wound                             0                    0             0
  Hardware                          0                    0             5
  Fracture                          0                    0             0
  Neurological                      0                    0             0
  Dislocation                       0                    0             0
```

```
  Loosening                       0                    0            2
  Thrombosis                      0                    0            0
  TumourSystemic                  0                    0            0
  TumourLocal                     0                    0            1
  InfectionSyst                   0                    0            0
  InfectionSuperficial            0                    1            1
  InfectionDeep                   1                    0            0
  Reoperation                     0                    1            3
  Amputation                      0                    0            1
  ImplantRemoval                  0                    0            1
  Mortality                       0                    0            0
  Censored                        0                    0            0
                    to
from                 Reoperation Amputation ImplantRemoval Mortality Censor
ed
  (s0)                          2          0              0         6
44
  Wound                         5          1              0         1
1
  Hardware                      3          0              0         0
5
  Fracture                      3          0              0         0
1
  Neurological                  1          0              0         0
8
  Dislocation                   9          0              0         0
1
  Loosening                     3          0              0         0
1
  Thrombosis                    0          0              0         1
1
  TumourSystemic                2          0              0         3
1
  TumourLocal                   2          2              0         1
4
  InfectionSyst                 0          0              0         1
1
  InfectionSuperficial          1          0              0         0
2
  InfectionDeep                11          2              1         0
1
  Reoperation                   0          1              0         1
19
  Amputation                    0          1              3         0
1
  ImplantRemoval                0          0              0         0
1
  Mortality                     0          0              0         0
0
  Censored                      0          0              0         0
```

```
0

Number of subjects with 0, 1, ... transitions to each state:
                      count
state                   0  1  2  3 4 5 6 7 10 12 17 18
  Wound                98  7  1  0 0 0 0 0  0  0  0  0
  Hardware             90 13  3  0 0 0 0 0  0  0  0  0
  Fracture            101  5  0  0 0 0 0 0  0  0  0  0
  Neurological         95 10  0  1 0 0 0 0  0  0  0  0
  Dislocation          99  4  1  2 0 0 0 0  0  0  0  0
  Loosening           100  6  0  0 0 0 0 0  0  0  0  0
  Thrombosis          103  3  0  0 0 0 0 0  0  0  0  0
  TumourSystemic      100  5  1  0 0 0 0 0  0  0  0  0
  TumourLocal          97  7  2  0 0 0 0 0  0  0  0  0
  InfectionSyst       104  2  0  0 0 0 0 0  0  0  0  0
  InfectionSuperficial 101  4  1  0 0 0 0 0  0  0  0  0
  InfectionDeep        93  8  4  0 1 0 0 0  0  0  0  0
  Reoperation          79 16  9  0 2 0 0 0  0  0  0  0
  Amputation          102  2  1  1 0 0 0 0  0  0  0  0
  ImplantRemoval      102  4  0  0 0 0 0 0  0  0  0  0
  Mortality            92 14  0  0 0 0 0 0  0  0  0  0
  Censored             14 92  0  0 0 0 0 0  0  0  0  0
  (any)                 0 50 25 12 5 4 3 3  1  1  1  1
```

#### 9.3 STROBE [12.3] Missingness evaluation and management

Missingess was assessed with visualisation and table functions in the *naniar* package (v1.1.0) [ (Tierney and Cook 2023) and compiled into figures using *patchwork* (v1.2.0.9000) [ (Pedersen 2023).

#### 9.4 STROBE [12.4] Analysis

##### 9.4.1 Diagnosis

The diagnosis text (see STROBE [8] for description) was processed using *tidytext* and integrated with the treatment record data for the index procedures. Stop words were removed and the text split into word tokens. The token length was calculated and abbreviations processed in the same manner as STROBE 12.2. The processed tokens were concatenated within record identifiers and integrated back into the index procedure table. The resulting dataframe was converted to a document-term matrix for topic modelling. Topic modelling was commenced with latent dirichlet allocation using the *ldatuning* package (v1.0.2 (Nikita 2020), stepping through possible topic numbers of 2 to 15, increasing by one per iteration. Two metrics were visually inspected (Cao et al. 2009; Deveaud, SanJuan, and Bellot 2014) from the resulting plot.

##### 9.4.2 Kaplan-Meier Analysis

To provide clinical guidance for postoperative expectations and to align with the broader literature, the analysis was repeated with Kaplan-Meier survival analysis (single event per record) for key adverse events. Data was arranged into wide format with each adverse event category organised into an indicator column and a duration column.

##### 9.4.3 Adverse Events and Multi-State Modelling

The analysis of adverse events and treatment/patient survival after arthroplasty remains a challenging endeavour, made more so by the complexities of ortho-oncology. Attempts have been made to standardize reporting of adverse outcomes after lower limb arthroplasty [citations], and also after endoprosthesis tumour surgery with the Henderson failure criteria [citation]. However, a key challenge of reporting incidence rates of these outcomes in a given sample is the variability in follow up from one patient to another in the same analytical sample. With variation in followup, the uni-dimensional estimate of incidence (number with condition/total available sample) leads to considerable underestimation of the true rate, since some cases have not yet reached sufficient followup to experience the event of interest. For this reason time-to-event (survival) analysis provides superior incidence estimates - however, there are additional aspects of the present analysis that preclude the use of traditional Kaplan-Meier analysis.

The first is that each patient can experience multiple adverse events after the index procedure (recurring events) which adds a element of dependency to the structure of the adverse event data (Thenmozhi et al. 2019), which is not accounted for in a KM curve. The second is that certain events (e.g. mortality or implant removal) preclude the appearance of subsequent adverse events. When these records are subsequently censored (removed from the pool available records) it can bias estimates of other events of interest upward to impossible values (Coemans et al. 2022). In the present dataset, where these elements exist simultaneously, traditional (simplistic) methods can lead to analytical decisions that remove a considerable amount of information from the dataset (e.g. analysis of first occurrence of any type) or biased estimates.

To address these issues within the analysis, the *survival* and *tidycmprsk* (v1.0.0)(Sjoberg and Fei 2023) packages were utilised to deploy a multi-state survival model (see Table 2) to estimate time-varying incidences of competing events such as;

- Mortality (competing)
- Implant removal (competing)
- Infection
- Dislocation - Instability
- Local tumour recurrence in tumour cases

The survival models were expressed in the form below, with each survival model was restricted to the subset of cases labelled by cohort (Tumour Pelvis or Reintervention Hip Arthroplasty).

```
CRModelTumour <- survfit2(Surv(DurationStart, DurationStop, Category) ~ 1,
                          data = ComplicMaster,
```

```
                          id = CombID,
                          subset = (RegistryCohortName == "TumourPelvis")
                          )

CRModelRHA <- survfit2(Surv(DurationStart, DurationStop, Category) ~ 1,
                       data = ComplicMaster,
                       id = CombID,
                       subset = (RegistryCohortName == "ReinterventionTHA")
                       )
```

##### 9.4.4 Henderson Failure

Records of treatment that had been transitioned to a subsequent record due to change in status aligned with a failure of treatment were categorised for the TumourPelvis cohort using the modified Henderson criteria for classification (Henderson et al. 2014).

### 10. STROBE [13] Participants

The initial export from the registry returned 1099 patient records and 1367 treatments. A total of 866 entries to the adverse events table of all types (including entries indicating no complications present) were retrieved.

#### 10.1 STROBE [13.1] Treatment selection

The procedures of interest were labelled with the manufacturer tag ("Ossis") for further retrieval. A total of 109 cases of all types were identified with 0 cases offered but refused and 3 planned but not implanted. The three cases for indication of pelvic tumour not implanted were abandoned due to;

- intraoperative blood loss; wound contamination and procedure duration
- intraoperative blood loss
- disease progression at time of surgery

The remaining 106 cases were the first use of the implant on a hemisphere of the pelvis, with 1 patient receiving sequential bilateral treatments for revision of total hip arthroplasties.

### 11. STROBE [14] Patient and record characteristics

Patient characteristics for cases receiving the implant of interest are summarised in Table 3 using the *gtsummary* package (v2.0.0) (Sjoberg et al. 2021).

Table 3 - Summary of patient characteristics cases receiving implant of interest (index procedure)

| **Characteristic** | **N** | **Overall** N = 106 | **95% CI**[1] | **ReinterventionTHA** N = 33 | **95% CI**[1] | **TumourPelvis** N = 73 | **95% CI**[1] |
|---|---|---|---|---|---|---|---|
| Age at Surgery, Mean (SD) | 106 | 55.0 (19.6) | 51 - 59 | 68.5 (9.8) | 65 - 72 | 48.8 (19.9) | 44 - 53 |
| Female, % (n) | 106 | 53% (56) | 43 - 63 | 70% (23) | 51 - 84 | 45% (33) | 34 - 57 |
| AffectedSide, % (n) | 106 | | | | | | |
| Axial | | 3.8% (4) | 1.2 - 9.9 | 0% (0) | 0.00 - 13 | 5.5% (4) | 1.8 - 14 |
| Left | | 50% (53) | 41 - 59 | 61% (20) | 42 - 77 | 45% (33) | 34 - 57 |
| Right | | 46% (49) | 37 - 56 | 39% (13) | 23 - 58 | 49% (36) | 38 - 61 |
| Surgeon, % (n) | 106 | | | | | | |
| A | | 54% (57) | 44 - 63 | 58% (19) | 39 - 74 | 52% (38) | 40 - 64 |
| B | | 41% (43) | 31 - 51 | 42% (14) | 26 - 61 | 40% (29) | 29 - 52 |
| C | | 1.9% (2) | 0.33 - 7.3 | 0% (0) | 0.00 - 13 | 2.7% (2) | 0.48 - 10 |
| D | | 3.8% (4) | 1.2 - 9.9 | 0% (0) | 0.00 - 13 | 5.5% (4) | 1.8 - 14 |

[1]CI = Confidence Interval

#### 11.1 STROBE [14.1] Pathology characteristics

A figure was created to assess the frequency of words used in the diagnosis description.

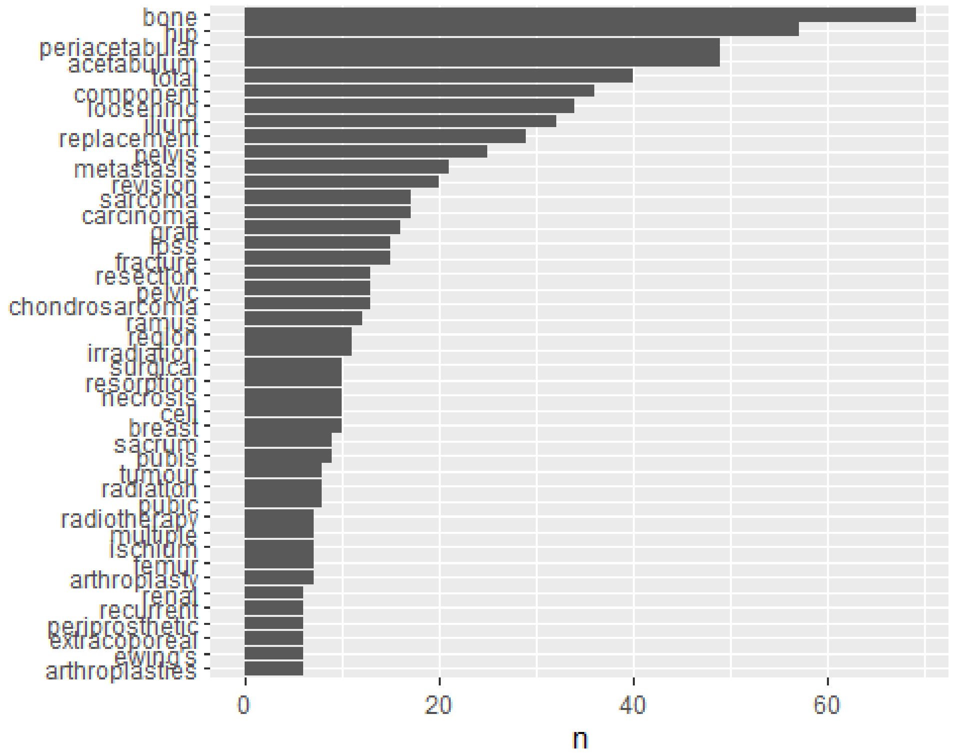

Figure 3: Word appearance in diagnosis text ordered by frequency.

```
fit models... done.
calculate metrics:
  CaoJuan2009... done.
  Deveaud2014... done.
```

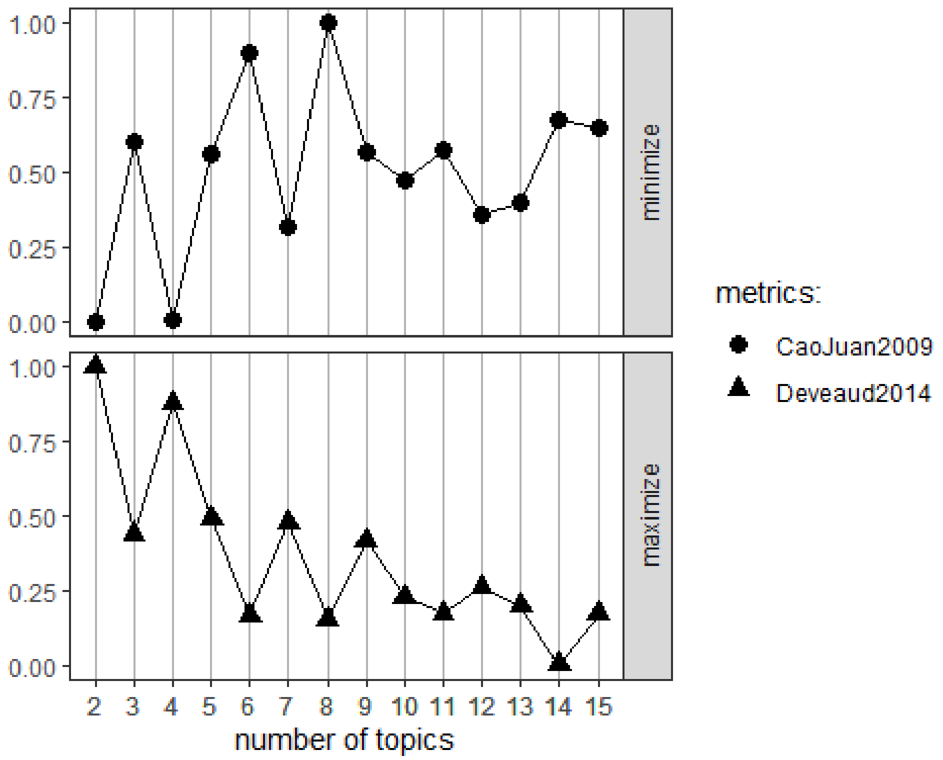

```
NULL
```

Figure 4: Topic metrics for Cao et al 2009 and Deveaud et al 2014 for models of 2 - 15 topics within diagnosis text

A final model with (n = 4) topics was computed using the *topicmodels* package (v0.2.16) (Grün and Hornik 2024). The Gibbs method was set with 500 iterations.

```
K = 4; V = 223; M = 106
Sampling 500 iterations!
Iteration 25 ...
Iteration 50 ...
Iteration 75 ...
Iteration 100 ...
Iteration 125 ...
Iteration 150 ...
Iteration 175 ...
Iteration 200 ...
Iteration 225 ...
Iteration 250 ...
Iteration 275 ...
Iteration 300 ...
```

```
Iteration 325 ...
Iteration 350 ...
Iteration 375 ...
Iteration 400 ...
Iteration 425 ...
Iteration 450 ...
Iteration 475 ...
Iteration 500 ...
Gibbs sampling completed!
```

Table 4: Proportion of topics observed in diagnosis text for three-dimensional printed custom pelvic implants

| | Proportion |
|---|---|
| bone periacetabular cell metastatic femoroacetabular | 28.4 |
| hip total component loosening replacement | 27.6 |
| ilium pelvis metastasis carcinoma sarcoma | 25.6 |
| acetabulum revision fracture ramus pelvic | 18.4 |

#### 11.2 STROBE [14.2] Management summary

#### 11.3 STROBE [14.3] Follow up

Table 5: Follow up duration of records included for analysis (regardless of outcome)

| **Characteristic** | **Overall** N = 106[1] | **ReinterventionTHA** N = 33[1] | **TumourPelvis** N = 73[1] |
|---|---|---|---|
| TreatDuration | 4.1 (2.4, 5.6) | 4.0 (2.4, 5.3) | 4.2 (2.4, 5.7) |

[1]Median (Q1, Q3)

The overall median follow up (IQR) of the series was 4.1 (2.4, 5.6) years, with a range of 0.6 - 10 years.

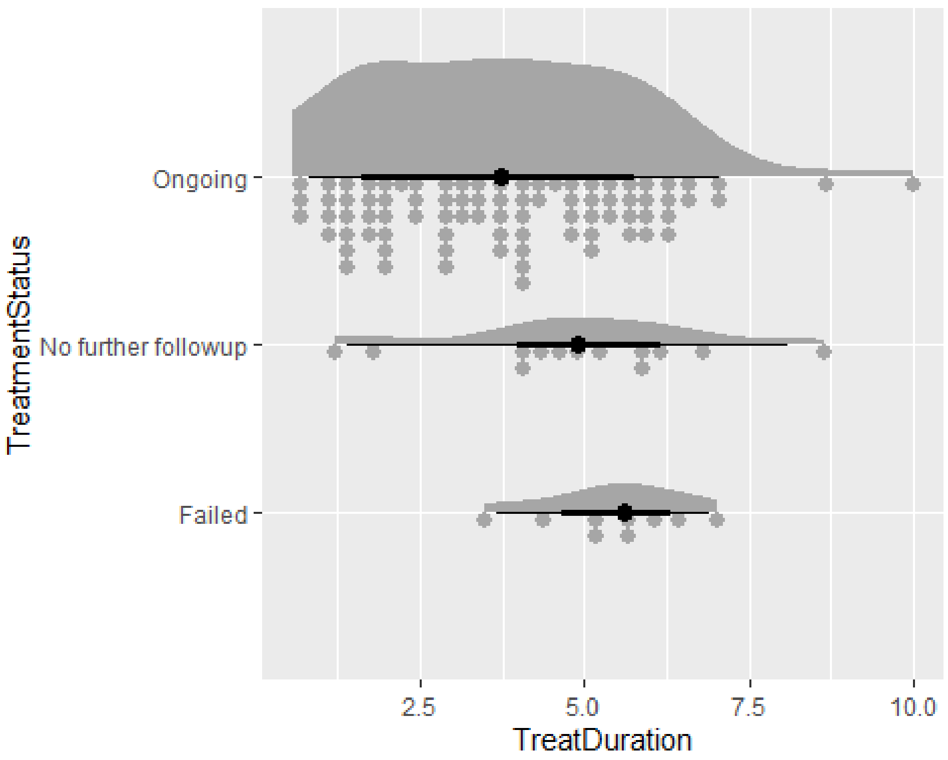

Figure 5: Follow up duration by treatment status

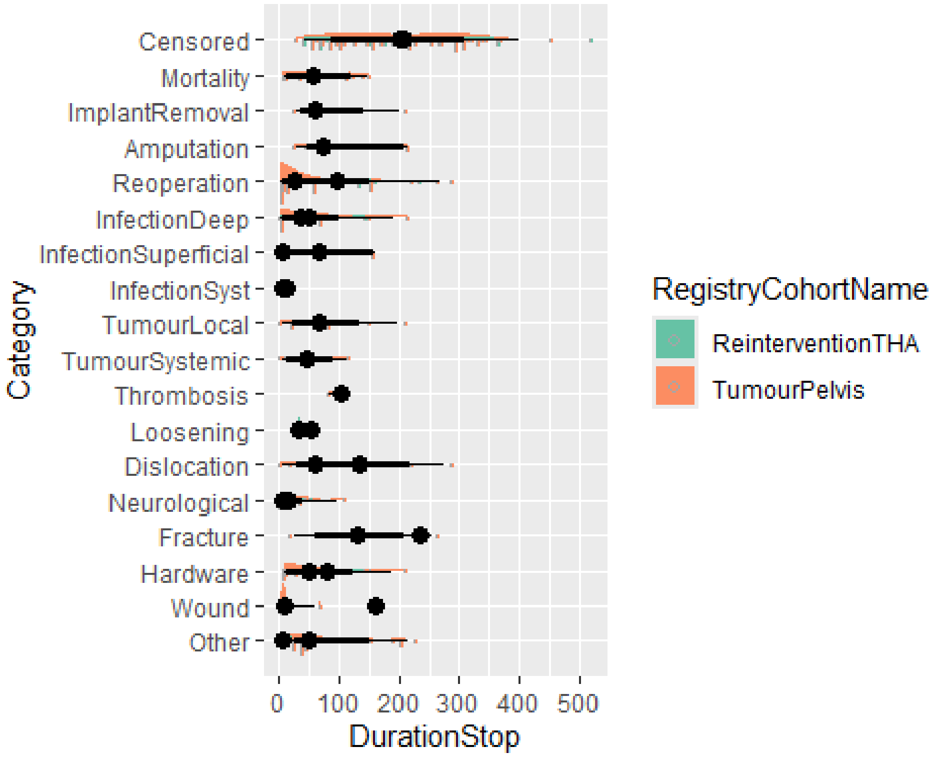

Figure 6: Follow up duration by adverse event category

#### 12. STROBE [15] Outcomes

There were 3 instances of intraoperative complications (4.1%, 95%CI 1.1 - 12.3) in the Tumour cohort of successful implantations. There was one major intraoperative haemorrhage, one confirmed positive margin and one possible positive margin detected by histopathology. All three cases remained alive with the implant in-situ at the end of the follow up period.

Table 6 - Summary of adverse events after receiving implant of interest (index procedure)

| **Characteristic** | **ReinterventionTHA** N = 33 | **95% CI**[1] | **TumourPelvis** N = 73 | **95% CI**[1] |
|---|---|---|---|---|
| Reoperation, n (%) | 8 (24) | 12 - 43 | 19 (26) | 17 - 38 |
| Hardware, n (%) | 2 (6.1) | 1.1 - 22 | 14 (19) | 11 - 30 |
| Dislocation, n (%) | 2 (6.1) | 1.1 - 22 | 5 (6.8) | 2.5 - 16 |
| InfectionSuperficial, n (%) | 1 (3.0) | 0.16 - 18 | 4 (5.5) | 1.8 - 14 |
| Other, n (%) | 2 (6.1) | 1.1 - 22 | 13 (18) | 10 - 29 |
| Mortality, n (%) | 0 (0) | 0.00 - 13 | 14 (19) | 11 - 30 |
| Wound, n (%) | 1 (3.0) | 0.16 - 18 | 7 (9.6) | 4.3 - 19 |
| Neurological, n (%) | 3 (9.1) | 2.4 - 25 | 8 (11) | 5.2 - 21 |
| TumourLocal, n (%) | 0 (0) | 0.00 - 13 | 9 (12) | 6.1 - 23 |
| InfectionDeep, n (%) | 4 (12) | 4.0 - 29 | 9 (12) | 6.1 - 23 |
| ImplantRemoval, n (%) | 0 (0) | 0.00 - 13 | 4 (5.5) | 1.8 - 14 |
| Amputation, n (%) | 0 (0) | 0.00 - 13 | 4 (5.5) | 1.8 - 14 |
| Fracture, n (%) | 1 (3.0) | 0.16 - 18 | 4 (5.5) | 1.8 - 14 |
| Loosening, n (%) | 2 (6.1) | 1.1 - 22 | 4 (5.5) | 1.8 - 14 |
| TumourSystemic, n (%) | 0 (0) | 0.00 - 13 | 6 (8.2) | 3.4 - 18 |
| InfectionSyst, n (%) | 1 (3.0) | 0.16 - 18 | 1 (1.4) | 0.07 - 8.4 |
| Thrombosis, n (%) | 0 (0) | 0.00 - 13 | 3 (4.1) | 1.1 - 12 |

[1]CI = Confidence Interval

There were no cases of mortality, tumour or thrombosis in the Reintervention THA cohort. Infections were observed (N = 4 (12)%, 95%CI 4.0 - 29%), as well as neurological issues - usually due to screw placement (N = 3 (9.1)%, 95%CI 2.4 - 25%).

The Tumour Pelvic cohort displayed mortality of N = 14 (19)% with relatively wide confidence intervals (95%CI 11 - 30)%, as well as infection (N = 9 (12)%, 95%CI 6.1 - 23%), local tumour recurrence (N = 9 (12)%, 95%CI 6.1 - 23%) and other hardware fixation issues (N = 14 (19)%, 95%CI 11 - 30%).

#### 13. STROBE [16] Main results

Overall, the implant retention rate for the series was (96.2%, 95%CI 90.1 - 98.8).

#### 13.1 Kaplan-Meier survival curves

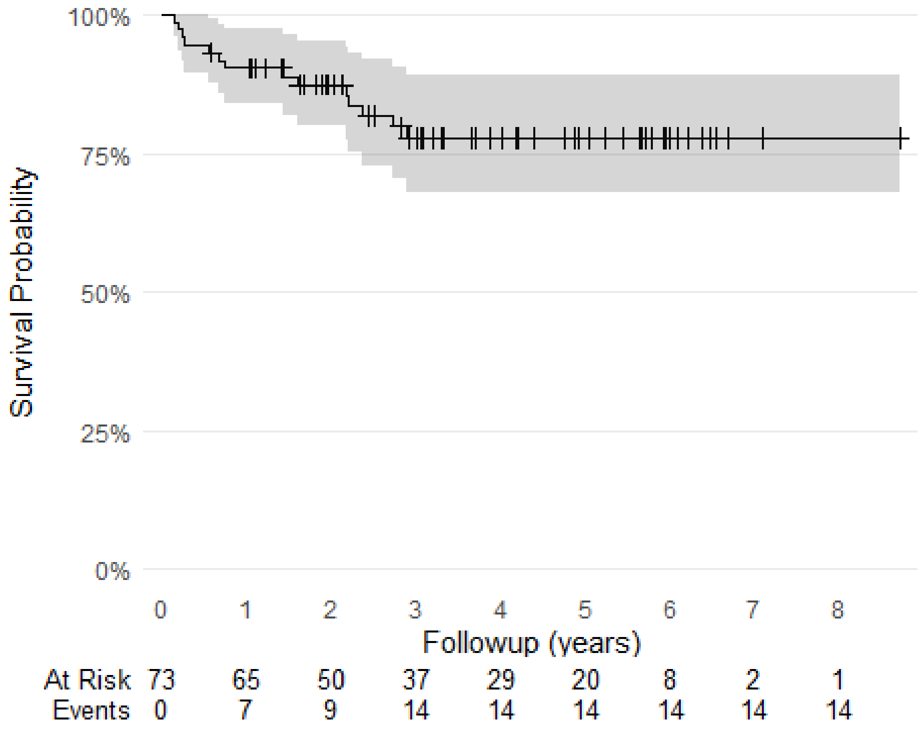

Figure 7: Kaplan-Meier survival curve for Mortality outcome in TumourPelvis cohort

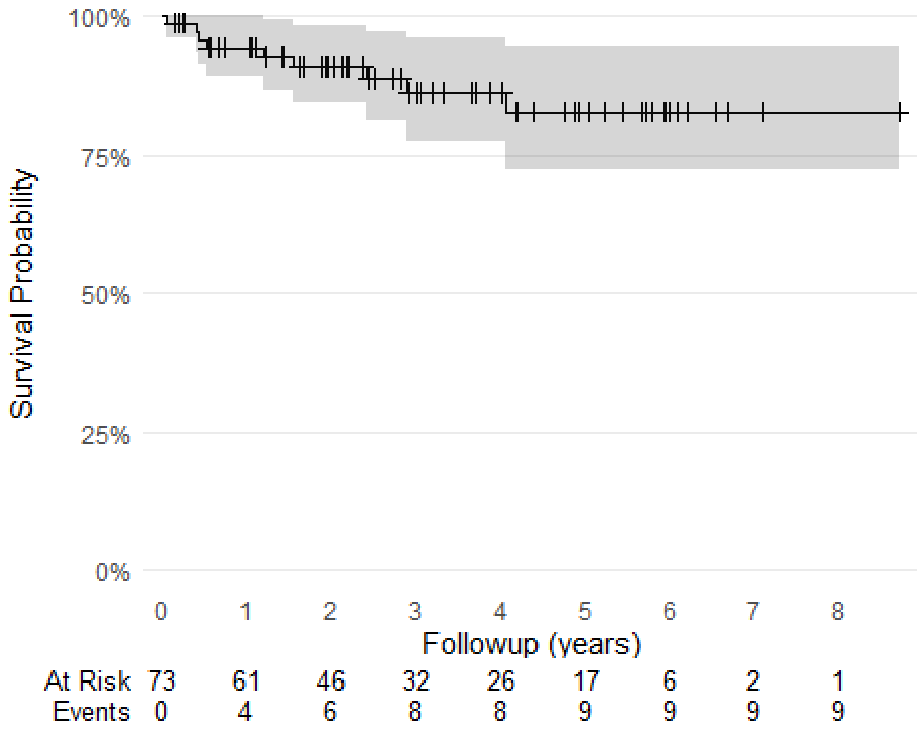

Figure 8: Kaplan-Meier survival curve for Local Tumour Recurrence in TumourPelvis cohort

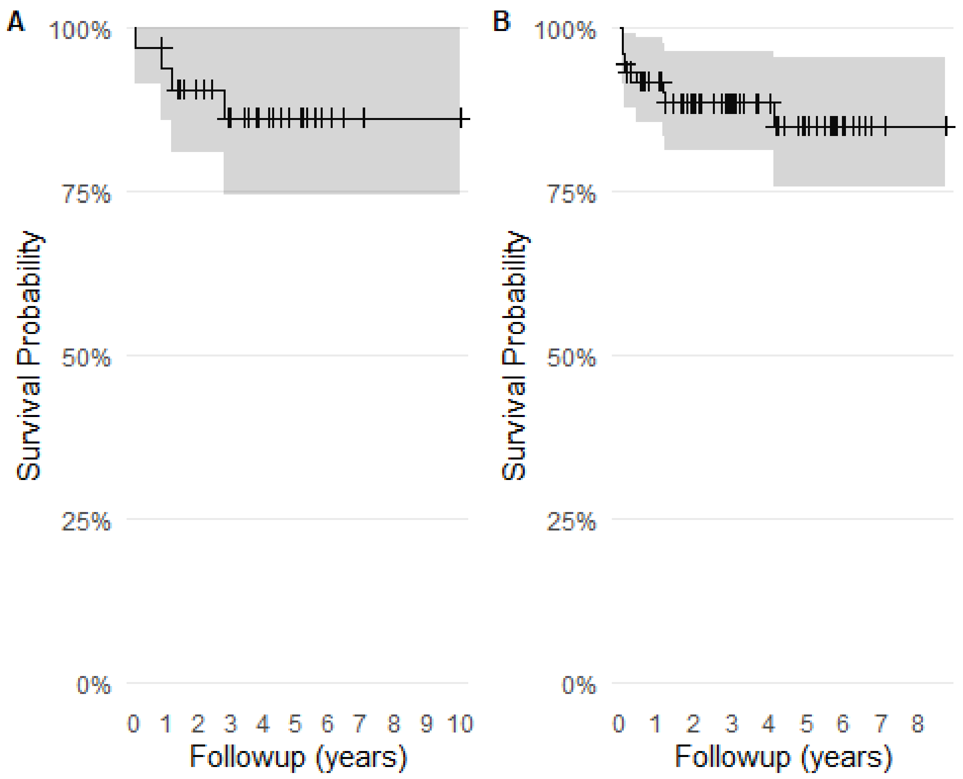

Figure 9: Kaplan-Meier survival curves for Infection in ReinterventionTHA (A) and TumourPelvis (B) cohorts

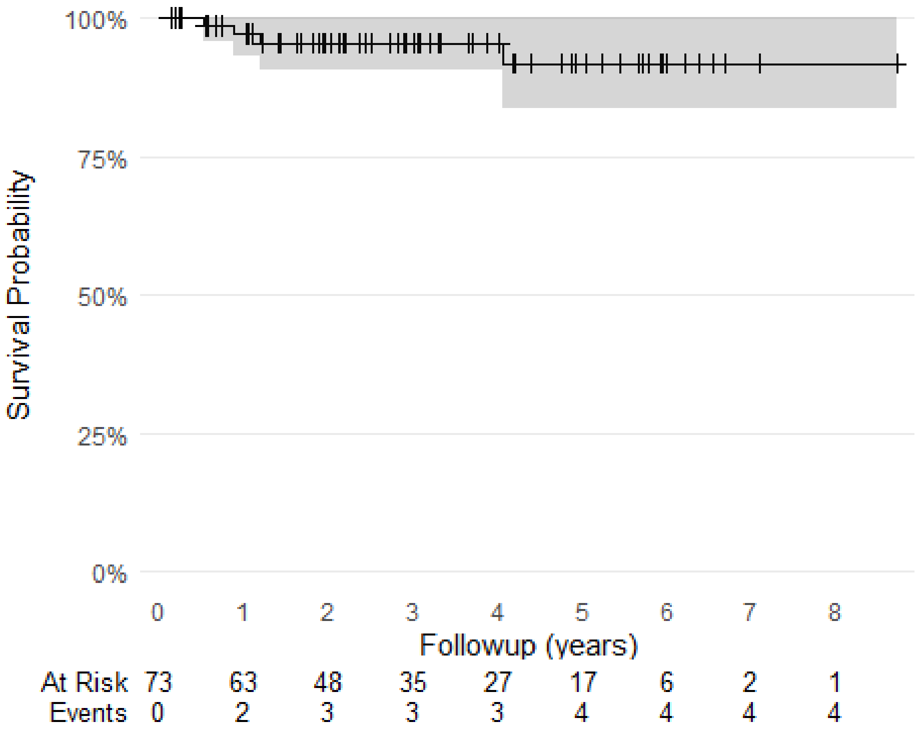

Figure 10: Kaplan-Meier survival curves for Amputation in TumourPelvis cohort

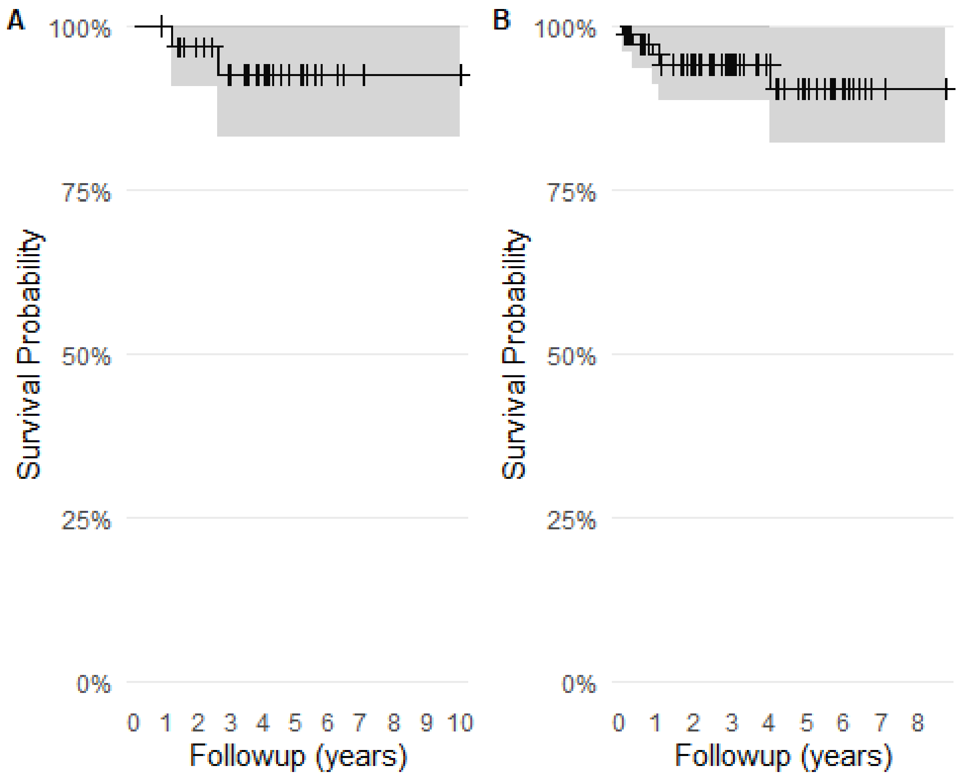

Figure 11: Kaplan-Meier survival curves for Dislocation in ReinterventionTHA (A) and TumourPelvis (B) cohorts

Table 9: Summary of Kaplan-Meier survival estimates for TumourPelvis cohort

| Characteristic | 0.5 Year | 1 Year | 2 Year | 5 Year |
| --- | --- | --- | --- | --- |
| Mortality | 95% (89% - 100%) | 90% (84% - 97%) | 87% (80% - 95%) | 78% (68% - 89%) |
| Local Recurrence | 96% (91% - 100%) | 94% (89% - 100%) | 91% (84% - 98%) | 83% (72% - 95%) |
| Periprosthetic Infection | 92% (86% - 98%) | 92% (86% - 98%) | 88% (81% - 96%) | 85% (76% - 95%) |
| Dislocation | 97% (93% - 100%) | 96% (91% - 100%) | 94% (89% - 100%) | 90% (82% - 100%) |
| Amputation and Implant Removal | 100% (100% - 100%) | 97% (93% - 100%) | 95% (90% - 100%) | 92% (84% - 100%) |

Table 10: Summary of Kaplan-Meier survival estimates for ReinterventionTHA cohort

| Characteristic | 0.5 Year | 1 Year | 2 Year | 5 Year |
| --- | --- | --- | --- | --- |
| Periprosthetic Infection | 97% (91% - 100%) | 94% (86% - 100%) | 91% (81% - 100%) | 86% (74% - 100%) |
| Dislocation | 100% (100% - 100%) | 100% (100% - 100%) | 97% (91% - 100%) | 93% (83% - 100%) |

#### 13.2 Multi-state survival model

Incidence rates were reviewed within the context of the multistate survival model. The cumulative incidences in the Tumour Pelvis cohort, when expressed at set follow up times, showed early peak incidence (<12months of surgery) for infection and local tumour recurrence and later incidence for dislocation and component loosening (Table 4). Cumulative mortality incidence also peaked at 20.6% by the 3 year followup.

The Reintervention THA cohort displayed a late peak in infection incidence (Table 5) at 2 years followup and an early peak in loosening (9 months).

Table 7: Summary of cumulative incidences of adverse events after custom pelvic implant for Tumour Pelvis

| | Wk14 | | | Wk38 | | | Wk52 | | | Wk104 | | | Wk156 | | |
|---|---|---|---|---|---|---|---|---|---|---|---|---|---|---|---|
| | CumIncid | CI Lower | CI Upper | CumIncid | CI Lower | CI Upper | CumIncid | CI Lower | CI Upper | CumIncid | CI Lower | CI Upper | CumIncid | CI Lower | CI Upper |
| (s0) | 69.9 | 60.1 | 81.2 | 56.2 | 45.9 | 68.8 | 52.1 | 41.8 | 64.9 | 30.1 | 21.3 | 42.7 | 20.5 | 13.1 | 32.3 |
| Wound | 2.7 | 0.7 | 10.7 | 1.4 | 0.2 | 9.6 | 1.4 | 0.2 | 9.6 | 1.4 | 0.2 | 9.6 | 1.4 | 0.2 | 9.6 |
| Hardware | 4.1 | 1.4 | 12.4 | 4.1 | 1.4 | 12.4 | 5.5 | 2.1 | 14.2 | 1.4 | 0.2 | 9.6 | 2.7 | 0.7 | 10.7 |
| Fracture | 0.0 | NA | NA | 1.4 | 0.2 | 9.6 | 1.4 | 0.2 | 9.6 | 2.7 | 0.7 | 10.7 | 0.0 | NA | NA |
| Neurological | 6.8 | 2.9 | 16.0 | 8.2 | 3.8 | 17.7 | 8.2 | 3.8 | 17.7 | 6.8 | 2.9 | 16.0 | 2.7 | 0.7 | 10.7 |
| Dislocation | 0.0 | NA | NA | 0.0 | NA | NA | 0.0 | NA | NA | 1.4 | 0.2 | 9.6 | 1.4 | 0.2 | 9.6 |
| Loosening | 0.0 | NA | NA | 0.0 | NA | NA | 0.0 | NA | NA | 1.4 | 0.2 | 9.6 | 1.4 | 0.2 | 9.6 |
| Thrombosis | 0.0 | NA | NA | 0.0 | NA | NA | 0.0 | NA | NA | 1.4 | 0.2 | 9.6 | 1.4 | 0.2 | 9.6 |
| TumourSystemic | 1.4 | 0.2 | 9.6 | 0.0 | NA | NA | 2.7 | 0.7 | 10.7 | 2.7 | 0.7 | 10.7 | 1.4 | 0.2 | 9.6 |
| TumourLocal | 0.0 | NA | NA | 1.4 | 0.2 | 9.6 | 1.4 | 0.2 | 9.6 | 2.7 | 0.7 | 10.7 | 4.1 | 1.4 | 12.4 |
| InfectionSyst | 0.0 | NA | NA | 0.0 | NA | NA | 0.0 | NA | NA | 0.0 | NA | NA | 0.0 | NA | NA |
| InfectionSuperficial | 1.4 | 0.2 | 9.6 | 0.0 | NA | NA | 0.0 | NA | NA | 1.4 | 0.2 | 9.6 | 2.7 | 0.7 | 10.7 |
| InfectionDeep | 0.0 | NA | NA | 1.4 | 0.2 | 9.6 | 0.0 | NA | NA | 0.0 | NA | NA | 0.0 | NA | NA |
| Reoperation | 8.2 | 3.8 | 17.7 | 15.1 | 8.7 | 26.0 | 13.7 | 7.7 | 24.4 | 12.3 | 6.7 | 22.7 | 9.6 | 4.7 | 19.4 |
| Amputation | 0.0 | NA | NA | 0.0 | NA | NA | 0.0 | NA | NA | 0.0 | NA | NA | 0.0 | NA | NA |
| ImplantRemoval | 0.0 | NA | NA | 1.4 | 0.2 | 9.6 | 2.7 | 0.7 | 10.7 | 2.7 | 0.7 | 10.7 | 1.4 | 0.2 | 9.6 |
| Mortality | 5.5 | 2.1 | 14.2 | 8.2 | 3.8 | 17.7 | 9.6 | 4.7 | 19.4 | 12.3 | 6.7 | 22.7 | 19.2 | 12.0 | 30.7 |
| Censored | 0.0 | NA | NA | 1.4 | 0.2 | 9.6 | 1.4 | 0.2 | 9.6 | 19.2 | 12.0 | 30.7 | 30.1 | 21.3 | 42.7 |

Table 8: Summary of cumulative incidences of adverse events after custom pelvic implant for Reintervention Total Hip Arthroplasty

| | Wk14 | | | Wk38 | | | Wk52 | | | Wk104 | | | Wk156 | | |
|---|---|---|---|---|---|---|---|---|---|---|---|---|---|---|---|
| | CumIncid | CI Lower | CI Upper | CumIncid | CI Lower | CI Upper | CumIncid | CI Lower | CI Upper | CumIncid | CI Lower | CI Upper | CumIncid | CI Lower | CI Upper |
| (s0) | 84.8 | 73.5 | 98.0 | 75.8 | 62.5 | 91.9 | 69.7 | 55.7 | 87.3 | 51.5 | 37.0 | 71.7 | 36.4 | 23.2 | 57.1 |
| Neurological | 6.1 | 1.6 | 23.2 | 6.1 | 1.6 | 23.2 | 9.1 | 3.1 | 26.7 | 6.1 | 1.6 | 23.2 | 6.1 | 1.6 | 23.2 |
| InfectionDeep | 3.0 | 0.4 | 20.9 | 3.0 | 0.4 | 20.9 | 3.0 | 0.4 | 20.9 | 3.0 | 0.4 | 20.9 | 3.0 | 0.4 | 20.9 |
| Reoperation | 0.0 | NA | NA | 6.1 | 1.6 | 23.2 | 9.1 | 3.1 | 26.7 | 15.2 | 6.8 | 34.0 | 21.2 | 11.0 | 40.9 |
| Censored | 0.0 | NA | NA | 0.0 | NA | NA | 6.1 | 1.6 | 23.2 | 21.2 | 11.0 | 40.9 | 33.3 | 20.6 | 54.0 |

The following figures illustrate the different incidence trajectories for adverse events within each cohort, when taking into account mortality and implant removal as competing risks.

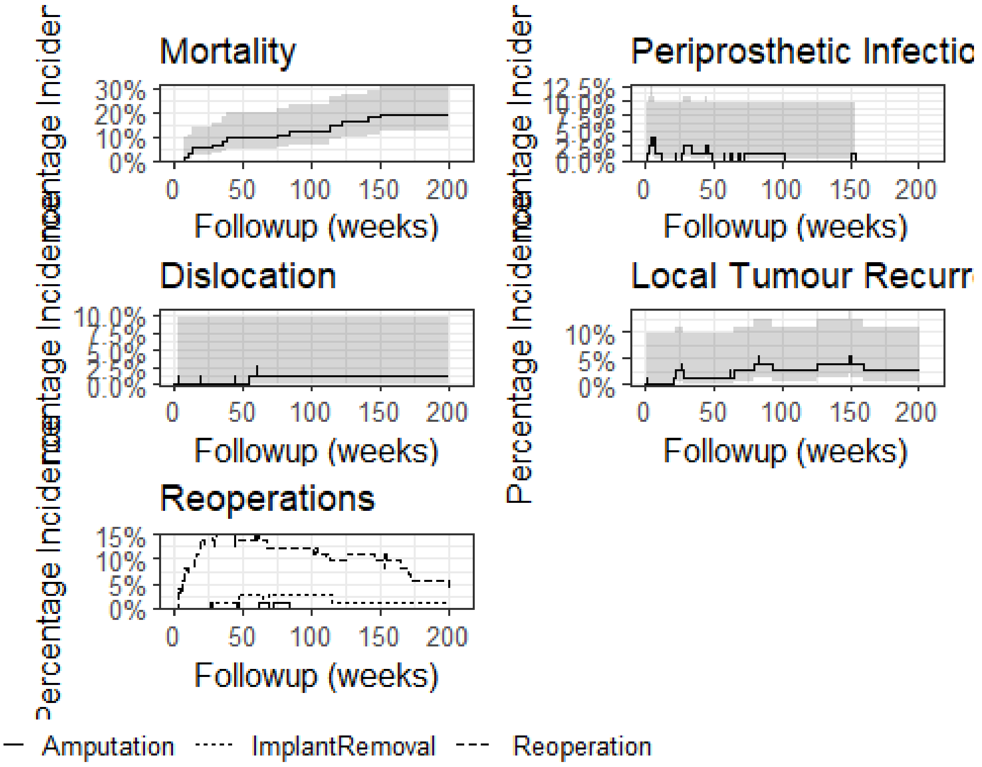

Figure 12: Cumulative incidence of adverse events after custom printed pelvic implant for pelvic tumour

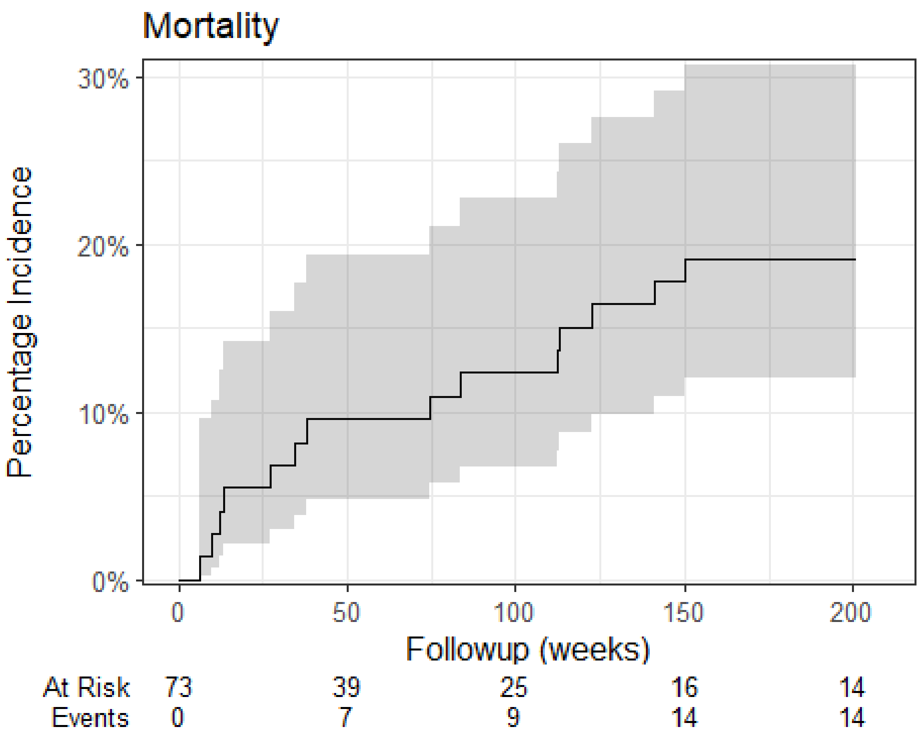

Mortality
30%
20%
10%
0%
Percentage Incidence
0
50
100
150
200
Followup (weeks)
At Risk 73 39 25 16 14
Events 0 7 9 14 14

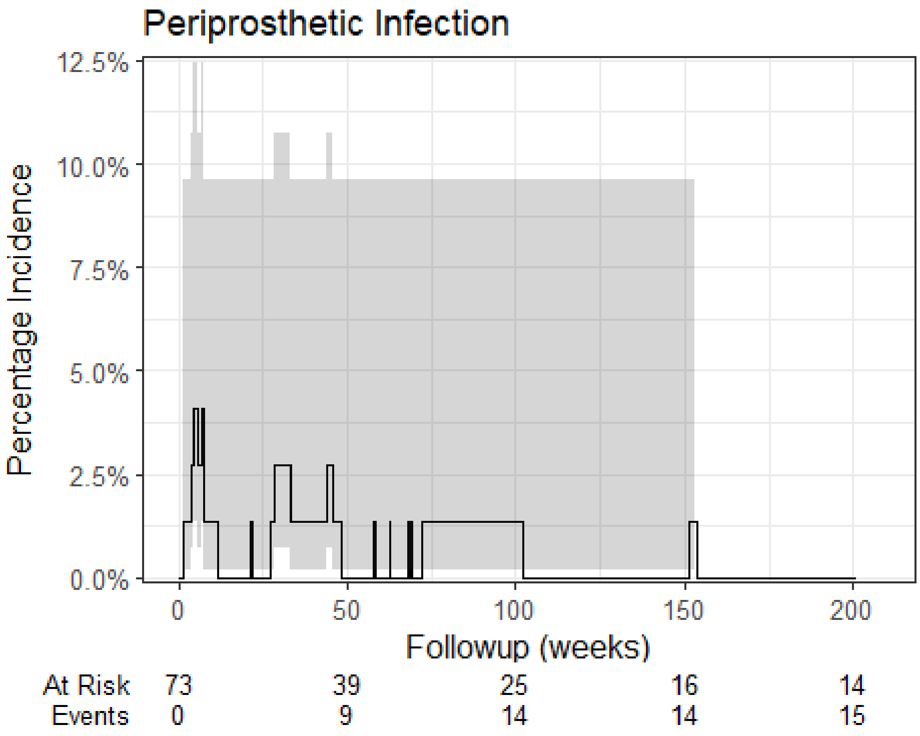

Periprosthetic Infection
12.5%
10.0%
7.5%
5.0%
2.5%
0.0%
Percentage Incidence
0
50
100
150
200
Followup (weeks)
At Risk 73 39 25 16 14
Events 0 9 14 14 15

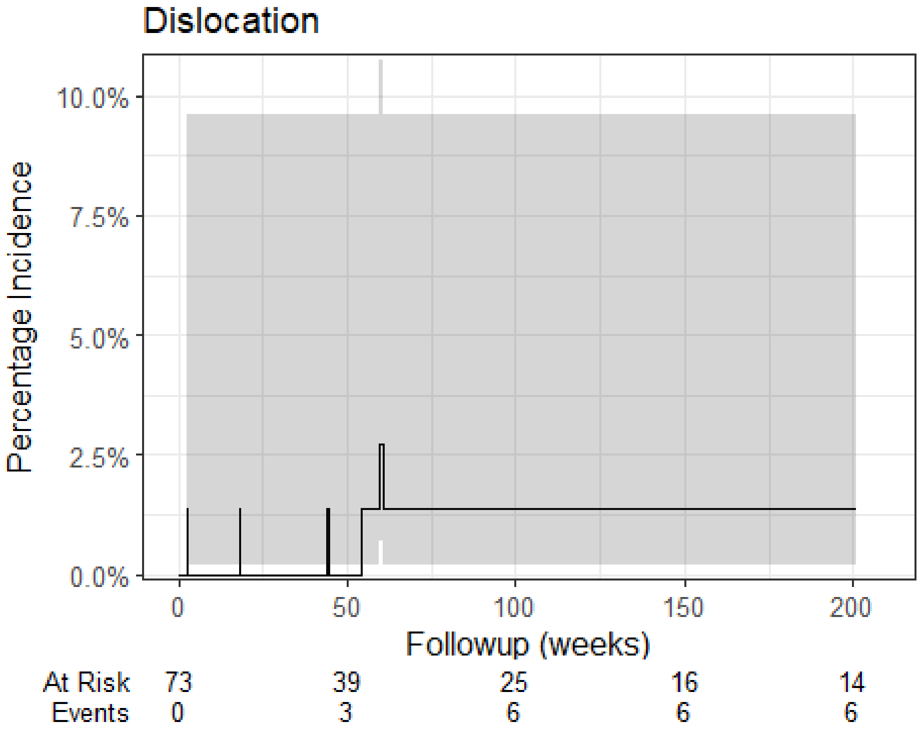

Dislocation
Percentage Incidence
10.0%
7.5%
5.0%
2.5%
0.0%
0
50
100
150
200
Followup (weeks)
At Risk 73 39 25 16 14
Events 0 3 6 6 6

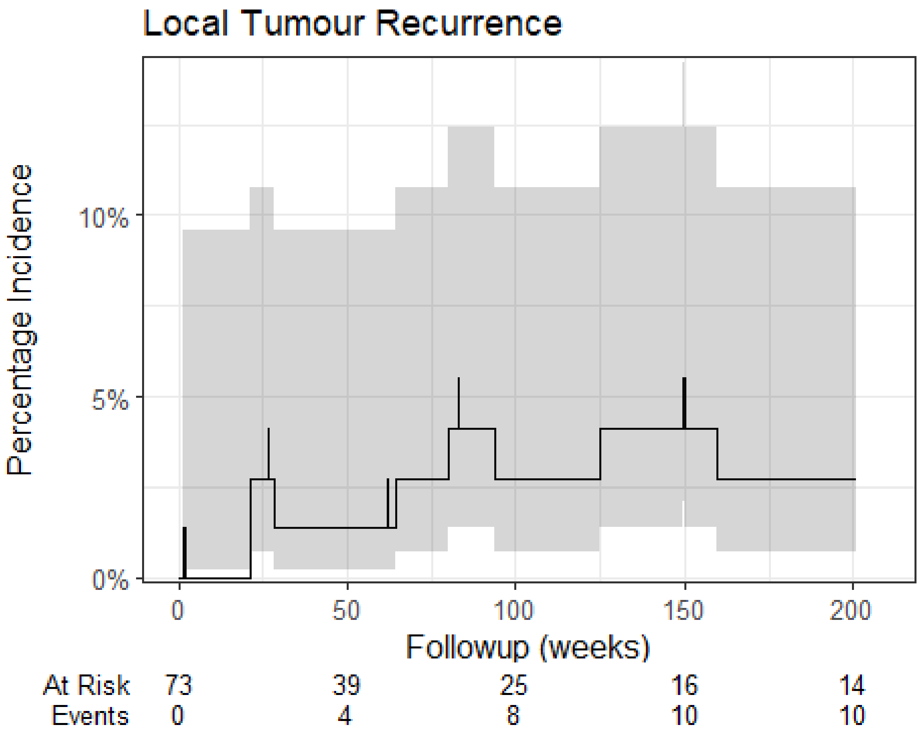

Local Tumour Recurrence
Percentage Incidence
10%
5%
0%
0
50
100
150
200
Followup (weeks)
At Risk 73 39 25 16 14
Events 0 4 8 10 10

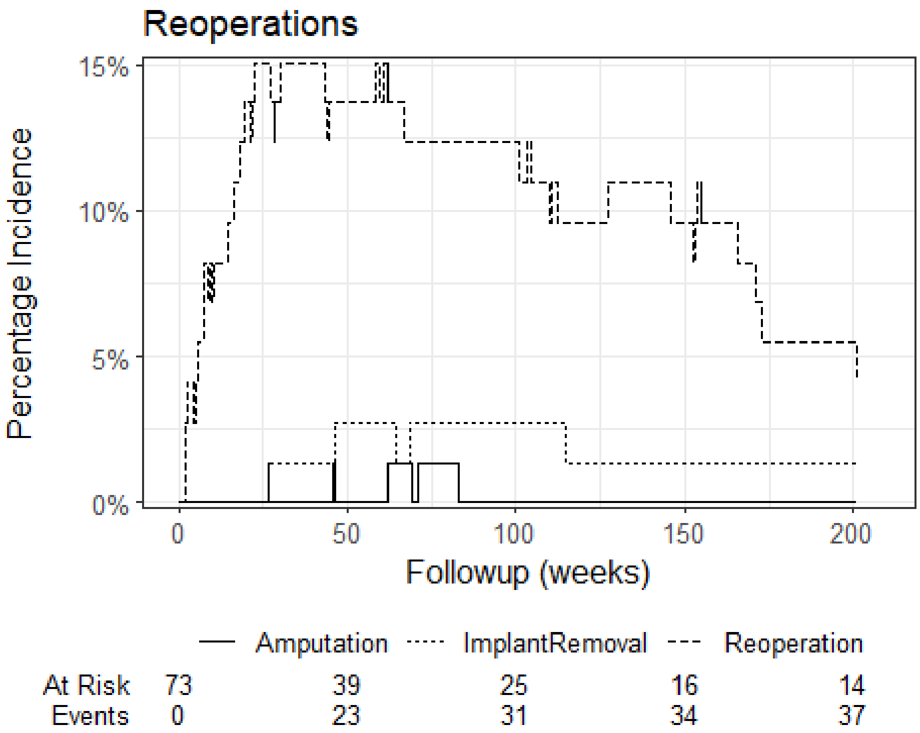
Reoperations
Percentage Incidence
15%
10%
5%
0%
0
50
100
150
200
Followup (weeks)
Amputation
ImplantRemoval
Reoperation
At Risk 73 39 25 16 14
Events 0 23 31 34 37

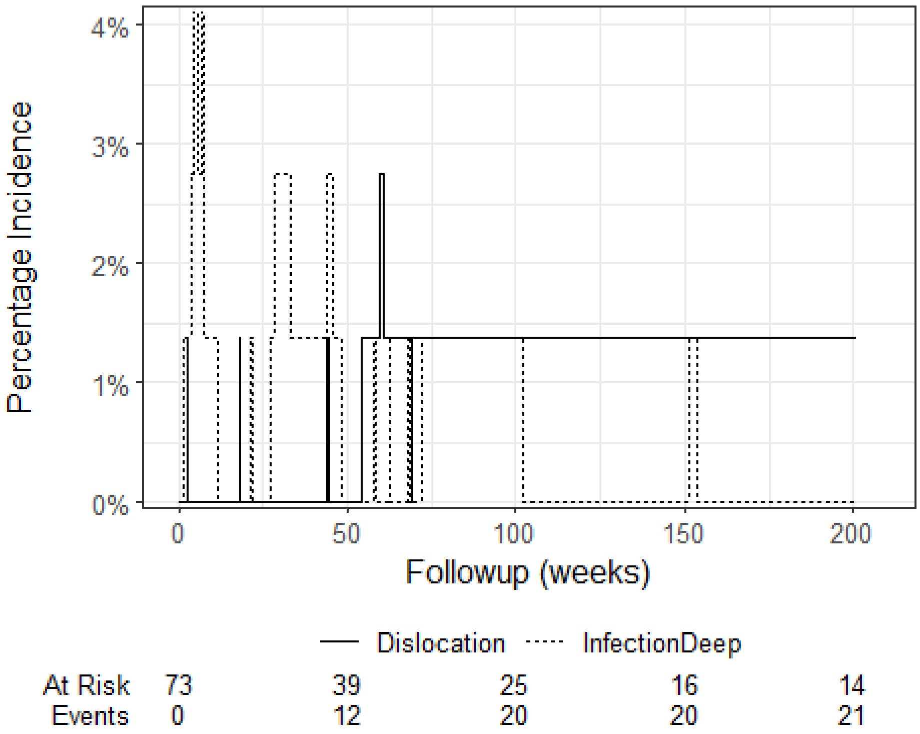
Percentage Incidence
4%
3%
2%
1%
0%
0
50
100
150
200
Followup (weeks)
Dislocation
InfectionDeep
At Risk 73 39 25 16 14
Events 0 12 20 20 21

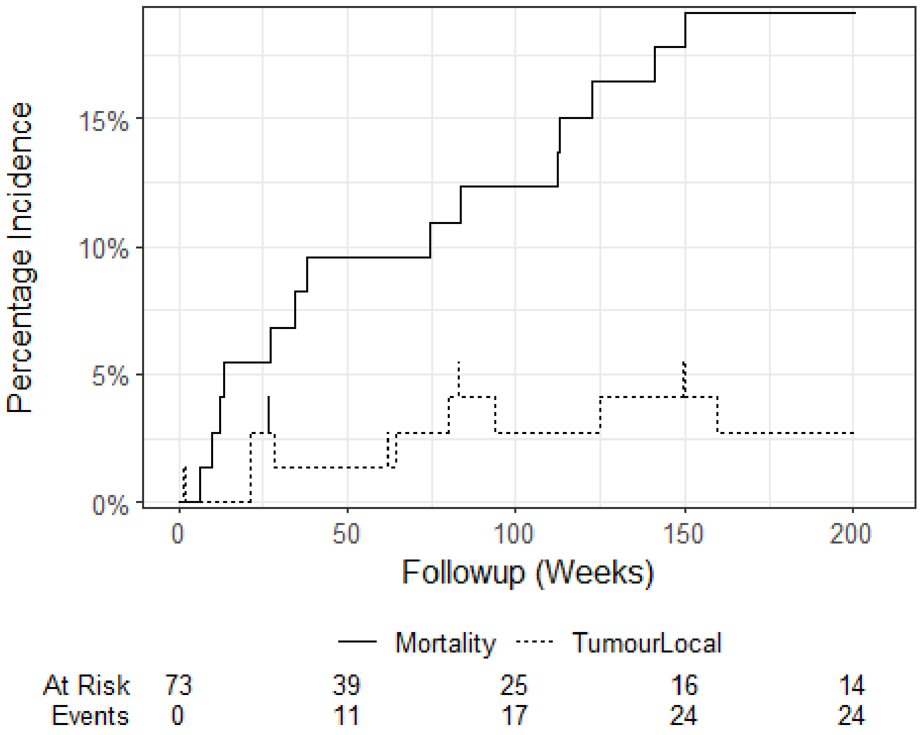

Figure 13: Risk of local tumour recurrence vs mortality after custom printed pelvic implant for pelvic tumour

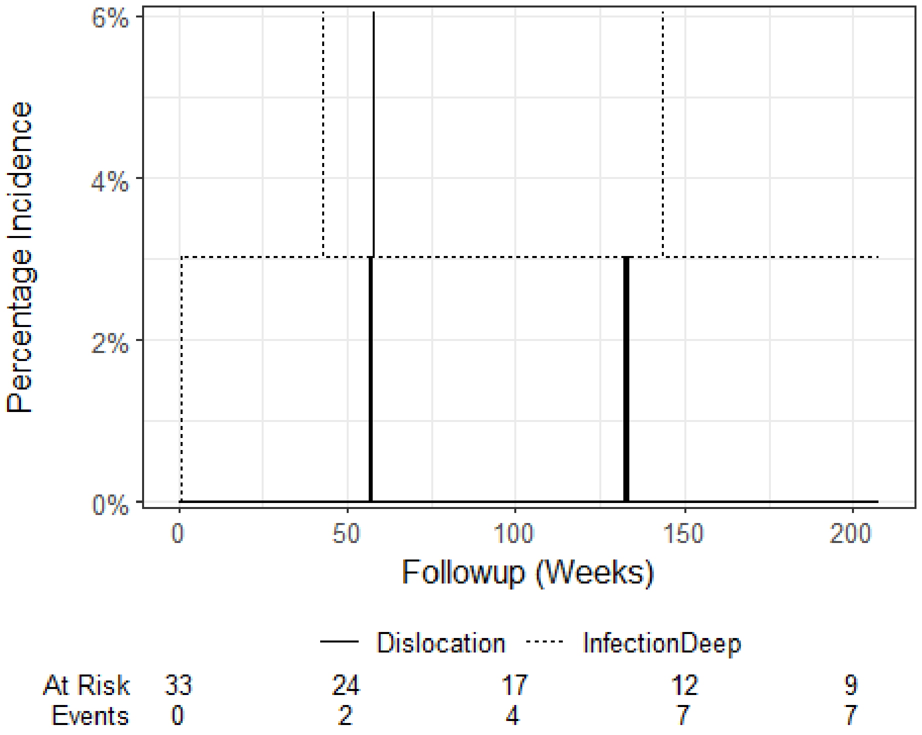

Figure 14: Cumulative risk of infection or dislocation after custom printed pelvic implant for reintervention total hip arthroplasty

##### 13.3 Henderson Failure Criteria

Treatment records considered "failed" were classified.

Nine cases were deemed to have failed leading to component revision or hip disarticulation. Of these;

- One recurrent hip dislocation was classified as Type 1
- One case of pelvic implant aseptic loosening was classified as Type 2
- Although cases of structural failure (periprosthetic fracture) were observed in the series, these events were all addressed with reoperations and were not considered failed.
- Two cases of infection (with one two-stage revision) were classified as Type 4
- Three were classified as Type 5 (tumour recurrence)
- Two cases of failure and THA revision could not be classified according to Henderson.
  - The first was a conversion of a proximal femoral modular component with a total femur construct due to loosening, unrelated to the pelvic implant.
  - The second was a first-occurrence hip dislocation where the acetabulum component was revised.

#### 14. STROBE [17] Sensitivity analyses

No sensitivity analyses were performed.

#### 15. Ethical Considerations:

##### 15.1 Informed Consent:

*Critically analyze the adequacy of procedures for obtaining informed consent from participants.*

A waiver of consent was granted to conduct chart review of the relevant patient records (see below).

##### 15.2 Ethical Approval:

*Evaluate whether the study received appropriate approval from an ethics committee or institutional review board.*

The study received HREC approval from the hospital HREC prior to commencement of chart review.

##### 15.3 Privacy and Confidentiality:

*Assess the adequacy of measures taken to protect participants' privacy and confidentiality. Beneficence and Non-maleficence: Critically evaluate any potential risks or benefits to participants and how they were addressed.*

Participant information was at risk of breach or leaking into the public domain. This risk was addressed by securing study materials using industry-standard methods to encrypt data at-rest within user-restricted data assets and in transit between the health service and the research team. Chart review was performed by approved persons with appropriate access and system credentials with training and experience on patient privacy and record management.

#### 16. Conflicts of Interest:

##### 16.1 STROBE [22] Funding:

*Assess the degree of transparency to which the source of funding and role of the funders have played in the study/article.*

The funding for this study was provided by the Department Research Fund. Some individuals responsible for administering the funds are co-authors of the study.

##### 16.2 Disclosure:

*Evaluate the transparency and completeness of disclosures regarding conflicts of interest among the study authors or sponsors.*

Funding for the establishment of the COMPRESSOR registry was received by Ossis Ltd. CS and ME declare institutional funding from Johnson and Johnson MedTech ANZ and MatOrtho Australia. RB declares institutional funding from Stryker South Pacific. RB and PS declare ownership of stocks in Ossis Ltd. MG and DF declare no conflict of interests.

##### 16.3 Potential for Biases:

*Critically assess whether conflicts of interest could have influenced the study design, analysis, or interpretation of results.*

Conflicts of interest could be perceived to influence the analysis or interpretation of results.

#### 17. Analysis References

Aden-Buie, Garrick. 2023. "Epoxy: String Interpolation for Documents, Reports and Apps." https://CRAN.R-project.org/package=epoxy.

Barbone, Jordan Mark, and Jan Marvin Garbuszus. 2024. "Openxlsx2: Read, Write and Edit 'Xlsx' Files." https://janmarvin.github.io/openxlsx2/.

Benchimol, Eric I., Liam Smeeth, Astrid Guttmann, Katie Harron, David Moher, Irene Petersen, Henrik T. Sørensen, Erik von Elm, and Sinéad M. Langan. 2015. "The REporting of Studies Conducted Using Observational Routinely-Collected Health Data (RECORD) Statement." *PLOS Medicine* 12 (10): e1001885. https://doi.org/10.1371/journal.pmed.1001885.

Bryan, Jennifer. 2023. "Googlesheets4: Access Google Sheets Using the Sheets API V4." https://CRAN.R-project.org/package=googlesheets4.

Cao, Juan, Tian Xia, Jintao Li, Yongdong Zhang, and Sheng Tang. 2009. "A Density-Based Method for Adaptive LDA Model Selection." *Neurocomputing* 72 (7-9): 1775–81. https://doi.org/10.1016/j.neucom.2008.06.011.

Coemans, Maarten, Geert Verbeke, Bernd Döhler, Caner Süsal, and Maarten Naesens. 2022. "Bias by Censoring for Competing Events in Survival Analysis." *BMJ*, September, e071349. https://doi.org/10.1136/bmj-2022-071349.

Davies, G. Matt, and Alan Gray. 2015. "Don't Let Spurious Accusations of Pseudoreplication Limit Our Ability to Learn from Natural Experiments (and Other Messy Kinds of Ecological Monitoring)." *Ecology and Evolution* 5 (22): 5295–5304. https://doi.org/10.1002/ece3.1782.

Deveaud, Romain, Eric SanJuan, and Patrice Bellot. 2014. "Accurate and Effective Latent Concept Modeling for Ad Hoc Information Retrieval." *Document Numérique* 17 (1): 61–84. https://doi.org/10.3166/dn.17.1.61-84.

Fellows, Ian. 2018. "Wordcloud: Word Clouds." https://CRAN.R-project.org/package=wordcloud.

Grün, Bettina, and Kurt Hornik. 2024. "Topicmodels: Topic Models." https://CRAN.R-project.org/package=topicmodels.

Hammer, Gaël P., Jean-Baptist du Prel, and Maria Blettner. 2009. "Avoiding Bias in Observational Studies." *Deutsches Ärzteblatt International*, October. https://doi.org/10.3238/arztebl.2009.0664.

Henderson, E. R., M. I. O'Connor, P. Ruggieri, R. Windhager, P. T. Funovics, C. L. Gibbons, W. Guo, F. J. Hornicek, H. T. Temple, and G. D. Letson. 2014. "Classification of Failure of Limb Salvage After Reconstructive Surgery for Bone Tumours." *The Bone & Joint Journal* 96-B (11): 1436–40. https://doi.org/10.1302/0301-620x.96b11.34747.

Iannone, Richard, Joe Cheng, Barret Schloerke, Ellis Hughes, Alexandra Lauer, and JooYoung Seo. 2024. "Gt: Easily Create Presentation-Ready Display Tables." https://CRAN.R-project.org/package=gt.

Lazic, Stanley E. 2010. "The Problem of Pseudoreplication in Neuroscientific Studies: Is It Affecting Your Analysis?" *BMC Neuroscience* 11 (1). https://doi.org/10.1186/1471-2202-11-5.

Man, Yvonne de, Yvonne Wieland-Jorna, Bart Torensma, Koos de Wit, Anneke L Francke, Mariska G Oosterveld-Vlug, and Robert A Verheij. 2023. "Opt-In and Opt-Out Consent Procedures for the Reuse of Routinely Recorded Health Data in Scientific Research and Their Consequences for Consent Rate and Consent Bias: Systematic Review." *Journal of Medical Internet Research* 25 (February): e42131. https://doi.org/10.2196/42131.

Nguyen, Van Thu, Mishelle Engleton, Mauricia Davison, Philippe Ravaud, Raphael Porcher, and Isabelle Boutron. 2021. "Risk of Bias in Observational Studies Using Routinely Collected Data of Comparative Effectiveness Research: A Meta-Research Study." *BMC Medicine* 19 (1). https://doi.org/10.1186/s12916-021-02151-w.

Nikita, Murzintcev. 2020. "Ldatuning: Tuning of the Latent Dirichlet Allocation Models Parameters." https://CRAN.R-project.org/package=ldatuning.

Pedersen, Thomas Lin. 2023. "Patchwork: The Composer of Plots." https://CRAN.R-project.org/package=patchwork.

Scholes, Corey, Kevin Eng, Meredith Harrison-Brown, Milad Ebrahimi, Graeme Brown, Stephen Gill, and Richard Page. 2023. "Patient Registry of Upper Limb Outcomes (PRULO): A Protocol for an Orthopaedic Clinical Quality Registry to Monitor Treatment Outcomes." *Journal of Surgical Protocols and Research Methodologies* 2023 (4). https://doi.org/10.1093/jsprm/snad014.

Silge, Julia, and David Robinson. 2016. "Tidytext: Text Mining and Analysis Using Tidy Data Principles in r" 1. https://doi.org/10.21105/joss.00037.

Sjoberg, Daniel D., and Teng Fei. 2023. "Tidycmprsk: Competing Risks Estimation." https://CRAN.R-project.org/package=tidycmprsk.

Sjoberg, Daniel D., Karissa Whiting, Michael Curry, Jessica A. Lavery, and Joseph Larmarange. 2021. "Reproducible Summary Tables with the Gtsummary Package" 13: 570–80. https://doi.org/10.32614/RJ-2021-053.

Thenmozhi, Mani, Visalakshi Jeyaseelan, Lakshmanan Jeyaseelan, Rita Isaac, and Rupa Vedantam. 2019. "Survival Analysis in Longitudinal Studies for Recurrent Events: Applications and Challenges." *Clinical Epidemiology and Global Health* 7 (2): 253–60. https://doi.org/10.1016/j.cegh.2019.01.013.

Therneau, Terry M. 2024. "A Package for Survival Analysis in r." https://CRAN.R-project.org/package=survival.

Tierney, Nicholas, and Dianne Cook. 2023. "Expanding Tidy Data Principles to Facilitate Missing Data Exploration, Visualization and Assessment of Imputations" 105. https://doi.org/10.18637/jss.v105.i07.

Tripepi, G., K. J. Jager, F. W. Dekker, C. Wanner, and C. Zoccali. 2008. "Bias in Clinical Research." *Kidney International* 73 (2): 148–53. https://doi.org/10.1038/sj.ki.5002648.

Vandenbroucke, Jan P, Erik von Elm, Douglas G Altman, Peter C Gøtzsche, Cynthia D Mulrow, Stuart J Pocock, Charles Poole, James J Schlesselman, and Matthias Egger. 2007. "Strengthening the Reporting of Observational Studies in Epidemiology (STROBE): Explanation and Elaboration." *PLoS Medicine* 4 (10): e297. https://doi.org/10.1371/journal.pmed.0040297.

Wickham, Hadley. 2016. "Ggplot2: Elegant Graphics for Data Analysis." https://ggplot2.tidyverse.org.

———. 2023. "Stringr: Simple, Consistent Wrappers for Common String Operations." https://CRAN.R-project.org/package=stringr.

Wickham, Hadley, Mara Averick, Jennifer Bryan, Winston Chang, Lucy D'Agostino McGowan, Romain François, Garrett Grolemund, et al. 2019. "Welcome to the Tidyverse" 4: 1686. https://doi.org/10.21105/joss.01686.

Wickham, Hadley, Romain François, Lionel Henry, Kirill Müller, and Davis Vaughan. 2023. "Dplyr: A Grammar of Data Manipulation." https://CRAN.R-project.org/package=dplyr.

Wickham, Hadley, and Lionel Henry. 2023. "Purrr: Functional Programming Tools." https://CRAN.R-project.org/package=purrr.

Xie, Yihui. 2024. "Knitr: A General-Purpose Package for Dynamic Report Generation in r." https://yihui.org/knitr/.
